# Biological aging markers in South African women with HIV and variable exposure to childhood maltreatment

**DOI:** 10.64898/2026.10.15.26365601

**Authors:** Kabelo Mabokane Simon Maloka, Georgina Spies, Stefan du Plessis, Sian Megan Joanna Hemmings, Scott Letendre, Soraya Seedat, Jacqueline Samantha Womersley

**Affiliations:** Department of Psychiatry, Faculty of Medicine and Health Sciences, Stellenbosch University, Cape Town, South Africa; SU/SAMRC Genomics of Brain Disorders Research Unit, Faculty of Medicine and Health Sciences, Stellenbosch University, Cape Town, South Africa; Department of Psychology, Faculty of Humanities, University of Cape Town, South Africa; Division of Infectious Diseases, Department of Medicine, University of California, San Diego, California, USA; Neuroscience Institute, Faculty of Health Sciences, University of Cape Town, South Africa; Department of Human Biology, Faculty of Health Sciences, University of Cape Town, South Africa

**Keywords:** HIV, Childhood Maltreatment, Biological Aging, Brain age, DNAmAge clocks

## Abstract

**Introduction:** HIV and childhood maltreatment (CM) are both chronic stressors that disproportionately affect women, with each contributing to accelerated biological aging. Whether biological aging markers are convergent and associate with HIV and CM is unclear.

**Methods:** We studied 238 women (122 with HIV), characterised for CM severity, and assessed biological aging using absolute telomere length (aTL), mitochondrial DNA copy number (mtDNAcn), magnetic resonance imaging-based brain-predicted age difference (MRI-BPAD), and DNA methylation age (DNAmAge) clocks. CM was measured with the Childhood Trauma Questionnaire–Short Form total score. We examined intercorrelations among biological aging markers to assess convergence and conducted linear regression models to test moderation by HIV status and CM severity.

**Results:** PWH had shorter aTL, lower MRI-BPAD, and higher Horvath age acceleration (HorvathAA), HannumAA, PhenoAA, and extrinsic epigenetic age acceleration (EEAA). CM had limited associations with biological aging markers. Absolute TL correlated weakly with mtDNAcn (*ρ* = 0.36), PhenoAA (*ρ* = -0.18), and EEAA (*ρ* = -0.18), while MRI-BPAD correlated weakly with PhenoAA (*r* = 0.20). DNAmAge clocks were moderately to strongly intercorrelated (*r* = 0.35 – 0.93). No robust two- or three-way moderation effects were observed after false discovery rate correction.

**Conclusion:** Our findings support a multidimensional pattern of biological aging, with substantial convergence among epigenetic aging measures but limited relationships across different markers, largely independent of HIV status and CM severity. Longitudinal approaches are needed to clarify how distinct biological aging markers relate to one another and to health outcomes over time in vulnerable populations.

## Introduction

HIV infection and childhood maltreatment (CM) are each chronic stressors that contribute to poor health outcomes and diminished quality of life (Bhutta et al., 2023; Olajide et al., 2024). These stressors are globally prevalent, and South Africa faces a disproportionate burden, particularly among women (Spies & Seedat, 2014; Steventon Roberts et al., 2022; UNAIDS, 2024). South Africa has one of the highest HIV prevalence rates worldwide, with women accounting for nearly 65% of people with HIV (PWH) in the country (Kharsany et al., 2018; UNAIDS, 2024). CM is also widespread, with high rates of physical, emotional, and sexual abuse reported among South African women (Gibbs et al., 2018; Myers et al., 2021; Steventon Roberts et al., 2022). Since both HIV and CM are highly prevalent, their co-occurrence and potential interactions are important to consider, including elucidating their individual and combined long-term biological consequences.

Following HIV infection, the virus crosses the blood-brain barrier, establishes reservoirs, and triggers chronic neuroinflammation (Huot et al., 2018; Saez-Cirion & Müller-Trutwin, 2019). These processes have been implicated in age-related health conditions such as neurocognitive disorders, cardiovascular diseases, and depression (Ghandakly et al., 2025; Gnoni, 2024; Thompson et al., 2024). Similarly, CM exerts lasting biological effects, including dysregulation of the hypothalamic-pituitary-adrenal axis and heightened inflammatory responses, which elevate the risk of age-related diseases (Nakama et al., 2023; Ridout et al., 2018). For example, high CM exposure may plausibly predispose individuals to inflammatory changes that are further exacerbated by HIV-related immune activation, compounding biological wear and tear (Renna et al., 2021; Wallace et al., 2020). Although several studies have documented dual exposure (Collins et al., 2023; Spies et al., 2016, 2017; Womersley et al., 2017, 2021), few have explicitly tested whether HIV and CM interact to accelerate biological aging. Addressing this gap is critical for identifying biomarkers and guiding interventions to mitigate long-term health consequences.

Biological aging markers, which reflect cumulative physiological wear and tear, are increasingly recognised as more informative than chronological age for assessing disease risk (Bell et al., 2019; Ferrucci et al., 2020; López-Otín et al., 2013). These include DNA methylation age (DNAmAge) clocks (Duan et al., 2022; Horvath, 2013), telomere length (TL) (Vostatek et al., 2023), mitochondrial DNA copy number (mtDNAcn) (Filograna et al., 2021; Vostatek et al., 2023), and magnetic resonance imaging-based brain-predicted age difference (MRI-BPAD) (Cole et al., 2018; Gampawar et al., 2022; Lee et al., 2022). Each marker has been individually associated with HIV-related outcomes and CM exposure (Cole et al., 2017; Fleming et al., 2025; Green et al., 2026; Horvath et al., 2018; Hulgan et al., 2019; Ma et al., 2026; Ridout et al., 2018; Womersley et al., 2021), yet it remains unclear whether they converge or diverge when considered together. Studies in people without HIV reported low to moderate correlations among these markers (Belsky et al., 2017; Kim et al., 2017; Li et al., 2020; Vetter et al., 2019; Zhang et al., 2018), with most correlations observed within marker types and not across different markers (Drewelies et al., 2025). Correlations among biological aging markers within PWH with CM exposure, particularly when multiple markers are assessed simultaneously, may therefore be informative in preliminarily understanding whether these markers demonstrate concordance and whether compounded stressors drive convergence or divergence in biological aging. No study has directly examined this question in populations facing the dual burden of HIV and CM, making it a critical gap that warrants future investigation.

To address this gap, we examined the interrelationships among nine widely studied biological aging markers, namely absolute TL (aTL), mtDNAcn, MRI-BPAD, and DNAmAge clocks (Horvath age acceleration [HorvathAA], HannumAA, PhenoAA, Horvath intrinsic epigenetic age acceleration [IEAA], Hannum IEAA, and extrinsic epigenetic age acceleration [EEAA]). We also assessed whether HIV status, CM severity, and their interaction moderated the associations between biological aging markers.

## Methods

### Participant recruitment and sample size composition

A total of 238 women (122 PWH, 116 controls without HIV) from a longitudinal study conducted between 2008 and 2015 were included in this cross-sectional analysis of baseline data. Participants were recruited from community health care facilities around Cape Town, South Africa. Sample sizes varied slightly across biological aging measures due to data availability but were generally balanced between groups. The largest sample was available for DNAmAge clocks (n = 231; 120 PWH, 111 controls), while the smallest overlap was for participants with all measures (n = 176; 90 PWH, 86 controls). Detailed sample size composition and overlap across measures are provided in **Supplementary Table S1**.

Recruitment procedures have been described previously (Spies et al., 2017). Participants aged 18-50 completed sociodemographic and neuropsychological assessments, and provided blood samples. Individuals with major psychiatric illness, substance use disorder, significant medical comorbidities, major prior head injury, probable dementia, opportunistic central nervous system infection or neoplasm, hepatitis B, or MRI contraindications were excluded. Controls tested HIV-negative through enzyme-linked immunosorbent assay. Immunovirological measures included CD4^+^ T cell counts (indicator of immune function and disease progression), CD8^+^ T cell counts (reflecting immune activation), the CD4:CD8 ratio (marker of immune balance), and viral load (measure of treatment response and ongoing viral replication).

### Assessment of CM and lifetime trauma exposure

CM was assessed using the 28-item Childhood Trauma Questionnaire – Short Form (CTQ-SF), which measures physical, emotional, and sexual abuse, as well as physical and emotional neglect before age 18 (Bernstein et al., 2003). Total scores were analysed continuously, with higher scores indicating greater CM. Internal consistency in this sample was good (α = 0.83). Lifetime trauma exposure was assessed using the Life Events Checklist (LEC), which evaluates exposure to 17 potentially traumatic events (Gray et al., 2004). Index events that were experienced or witnessed by participants were summed, with higher scores indicating greater trauma exposure (α = 0.77).

### MRI acquisition

T1-weighted MRI scans were acquired on a Siemens MAGNETOM Allegra 3T scanner (Erlangen, Germany) using either a magnetisation-prepared rapid gradient-echo (MPRAGE) sequence (n = 102) (repetition time [TR] = 2080 ms; voxel size = 1.3 x 1.0 x 1.3 mm; field of view [FOV] = 256 mm; echo time [TE] = 1.53 ms; inversion time [TI] = 1100 ms; flip angle = 7°) or a multi-echo MPRAGE sequence (n = 100) (TR = 2530 ms; TE1 = 1.53 ms, TE2 = 3.21 ms, TE3 = 4.89 ms, TE4 = 6.57 ms, flip angle = 7°, FOV = 256 mm, 128 slices, 1 mm isotropic voxel size).

### Biological aging measures

#### DNA methylation age clocks

Whole-blood DNA quality and purity were assessed using the Qubit 4 fluorometer (Invitrogen, Waltham, MA) or NanoDrop One spectrophotometer (Thermo Fisher Scientific, Waltham, MA). DNA samples were normalised to 350 ng total DNA and diluted to a concentration of 6 ng/µL before methylation profiling. DNA methylation profiling was performed at the University of Minnesota Genomics Center using the Infinium MethylationEPIC v2 BeadChip (Illumina, San Diego, CA). Quality-controlled DNA methylation data were generated using the *ENmix* pipeline in R (Xu et al., 2021), which included quality assessment and filtering of low-quality probes, samples, and individual methylation measurements based on detection P values, bead counts, bisulfite-conversion intensity, and outlier detection. The resulting beta values were submitted to the online DNAmAge calculator (https://dnamage.clockfoundation.org) to generate Horvath, Hannum, PhenoAge, Horvath IEAA, Hannum IEAA, and EEAA estimates. Age acceleration measures were calculated as the discrepancy between DNAmAge and chronological age.

#### Absolute telomere length and mitochondrial DNA copy number

DNA extracted from whole blood was stored at −80 °C until analysis. Absolute TL and mtDNAcn were quantified simultaneously using the absolute human TL and mtDNAcn dual quantification quantitative polymerase chain reaction (qPCR) assay kit (ScienCell Research Laboratories, Carlsbad, CA; Catalog number 8958) on a QuantStudio 6 Pro Real-Time PCR System (Thermo Fisher Scientific). Samples were analysed in triplicate according to the manufacturer’s protocols. Absolute TL was expressed as kilobases per chromosome end, and mtDNAcn was expressed as mitochondrial DNA copies per diploid cell.

#### MRI brain-predicted age

MRI-BPAD was estimated using the brainageR pipeline, which applies Gaussian process regression to voxel-wise tissue maps derived from SPM12 preprocessing (Clausen et al., 2022; Cole et al., 2017). The model was trained on 3377 healthy adults aged 18 - 92 years and has been validated across multiple cohorts (Biondo et al., 2022; Boyle et al., 2021; Dörfel et al., 2023; Hendrikse et al., 2025). MRI-BPAD was calculated as the brain-predicted age minus chronological age. Processing was performed on the Centre for High-Performance Computing server (https://www.chpc.ac.za/). The brainageR code is openly available (https://github.com/jamescole/brainageR).

### Model and assay validation

BrainageR performance was assessed using the mean absolute error (MAE), root mean squared error (RMSE), Pearson correlation between brain-predicted age and chronological age, and variance explained (Han et al., 2021). Performance was also assessed separately in PWH and controls. DNAmAge clocks were validated using Pearson correlations with chronological age. Absolute TL and mtDNAcn qPCR assay performance was assessed using melting curve plots and reference DNA two-fold serial dilution.

### Statistical analysis

Analyses were conducted in R 4.3.3 and figures were generated using *ggplot2* (R Core Team, 2021). Continuous variables were assessed for normality using Shapiro–Wilk tests and histograms. Group differences were evaluated using Mann–Whitney U, Welch’s t-tests, or chi-square tests as appropriate. Effect sizes were calculated using Cohen’s *d* for normally distributed variables and rank-biserial correlation for non-normal variables. Absolute TL and mtDNAcn were log-transformed and winsorised at the 95th percentile (Alegría-Torres et al., 2016; Chong et al., 2022). Associations between aging markers and CTQ-SF scores were examined using partial Spearman correlations. Partial Pearson correlations were applied when both variables were normally distributed, whereas partial Spearman correlations were used when at least one variable was non-normal. Moderation analyses employed linear regression models. Covariates were marker-specific and included age, age-squared (to account for the non-linear association between brain-predicted age and chronological age; Cole & Franke, 2017), MRI protocol type (MRI-BPAD models), plate number (aTL and mtDNAcn), and LEC scores (CTQ-SF analyses). False discovery rate (FDR) correction was applied to correlation and moderation analyses, with adjusted *p*-values < 0.05 considered statistically significant.

## Results

### Demographic and clinical characteristics

Participant sociodemographic and clinical characteristics are summarised in **Table 1**. The sample comprised 238 women with data available for at least one biological aging marker (122 PWH). PWH were significantly older than controls (*W* = 4247.0, *p* < 0.001) and had lower educational attainment (*ꭓ*^2^(2) = 4.7, *p* = 0.031), whereas employment status, ethnicity, and MRI protocol type did not differ between groups. CM total score and all CTQ-SF subtypes were higher in PWH than controls, while lifetime traumatic event exposure did not differ.

**Table 1.** Comparison of sociodemographic and clinical variables between PWH and controls without HIV.

| Characteristics | Total<br>N = 238 | PWH<br>n = 122 | Controls<br>n = 116 | Statistic |
| --- | --- | --- | --- | --- |
| Age, median (IQR) | 29.00 (25.00 – 36.00) | 33.00 (28.00 – 38.00) | 26.00 (22.00 – 33.00) | $W = 4247.00, p < 0.001^{***}$ |
| Education (≥ grade 9), n (%) | 207 (86.97) | 100 (81.97) | 107 (92.24) | $\chi^2(2) = 4.67, p = 0.031^*$ |
| Employed, n (%) | 70 (29.41) | 40 (32.79) | 30 (25.86) | $\chi^2(2) = 1.06, p = 0.303$ |
| Self-identified ethnicity, n (%) | | | | $\chi^2(2) = 0.21, p = 0.650$ |
| Black African | 234 (98.32) | 119 (97.54) | 115 (99.14) |  |
| South African Coloured<sup>1</sup> | 4 (1.68) | 3 (2.46) | 1 (0.86) |  |
| CTQ-SF total score, median (IQR) | 44.00 (31.00 – 58.75) | 52.50 (38.00 – 65.50) | 35.00 (27.75 – 47.00) | $W = 3749.50, p < 0.001^{***}$ |
| Emotional abuse, median (IQR) | 9.00 (5.00 – 15.00) | 12.00 (6.25 – 18.00) | 8.00 (5.00 – 11.25) | $W = 5192.50, p < 0.001^{***}$ |
| Emotional neglect, median (IQR) | 10.00 (6.00 – 15.00) | 12.50 (8.00 – 16.00) | 7.00 (5.00 – 11.00) | $W = 4050.50, p < 0.001^{***}$ |
| Physical abuse, median (IQR) | 5.00 (5.00 – 11.00) | 8.00 (5.00 – 13.75) | 5.00 (5.00 – 6.00) | $W = 4478.50, p < 0.001^{***}$ |
| Physical neglect, median (IQR) | 9.00 (6.00 – 12.00) | 10.00 (7.25 – 13.75) | 7.00 (5.00 – 9.00) | $W = 4302.50, p < 0.001^{***}$ |
| Sexual abuse, median (IQR) | 5.00 (5.00 – 7.00) | 5.00 (5.00 – 8.75) | 5.00 (5.00 – 5.00) | $W = 5540.50, p < 0.001^{***}$ |
| LEC score, median (IQR) | 5.00 (2.25 – 7.00) | 5.00 (3.00 – 7.00) | 4.00 (2.00 – 6.00) | $W = 6182.50, p = 0.091$ |
| MRI protocol type, n (%)<sup>2</sup> | | | | $\chi^2(2) = 0.00, p = 1.000$ |
| MPRAGE | 102 (50.50) | 53 (50.50) | 49 (50.50) |  |
| ME-MPRAGE | 100 (49.50) | 52 (49.50) | 48 (49.50) |  |
| Antiretroviral treatment, n (%) | - | 51 (41.80) | - | - |
| Log<sub>10</sub> VL, median (IQR) | - | 3.27 (1.64 – 4.31) | - | - |
| CD4 count, median (IQR) | - | 383.00 (252.00 – 591.75) | - | - |
| CD8 count, median (IQR)<sup>3</sup> | - | 956.00 (710.00 – 1089.25) | – | – |
| CD4:CD8 ratio, median (IQR) | - | 0.47 (0.31 – 0.65) | – | – |

**Abbreviations:** Childhood Trauma Questionnaire-Short Form (CTQ-SF), cluster of differentiation 4 (CD4), cluster of differentiation 8 (CD8), interquartile range (IQR), Life Events Checklist (LEC), magnetic resonance imaging (MRI), magnetisation-prepared rapid gradient-echo (MPRAGE), multi-echo magnetisation-prepared rapid gradient-echo (ME-MPRAGE), people with HIV (PWH), viral load (VL).
<sup>1</sup> “Coloured” is the official census category in South Africa (Statistics South Africa, 2025). The term is used here in its technical demographic sense and is not intended in a derogatory manner.
<sup>2</sup> MRI data were available for a subset of the total sample ( $n = 202$ )
<sup>3</sup> CD8 data were available for a subset of PWH ( $n = 44$ ).
**Significance:** \* $p < 0.05$ , \*\* $p < 0.01$ , \*\*\* $p < 0.001$

### Biological aging markers and chronological age

Biological aging markers were validated against chronological age (**Figure 1**). Assay performance metrics for aTL and mtDNAcn are presented in **Supplementary Figures S1 and S2**. Log-winsorised aTL was not associated with chronological age, whereas log-winsorised mtDNAcn showed a very weak correlation (*r* = 0.15, *p* = 0.031). Among participants with MRI data, the MRI-based brain-predicted age was moderately associated with chronological age (*r* = 0.50, *p* < 0.001), with MAE of 5.59 years and RMSE of 7.22 years. These error values are consistent with prior brain age studies, underscoring the validity of our model (Clausen et al., 2022; Han et al., 2021; Hendrikse et al., 2025). In the DNAmAge dataset, chronological age was strongly correlated with Horvath, Hannum, and PhenoAge clocks (*r* = 0.77–0.83, all *p* < 0.001).

**Figure 1.**
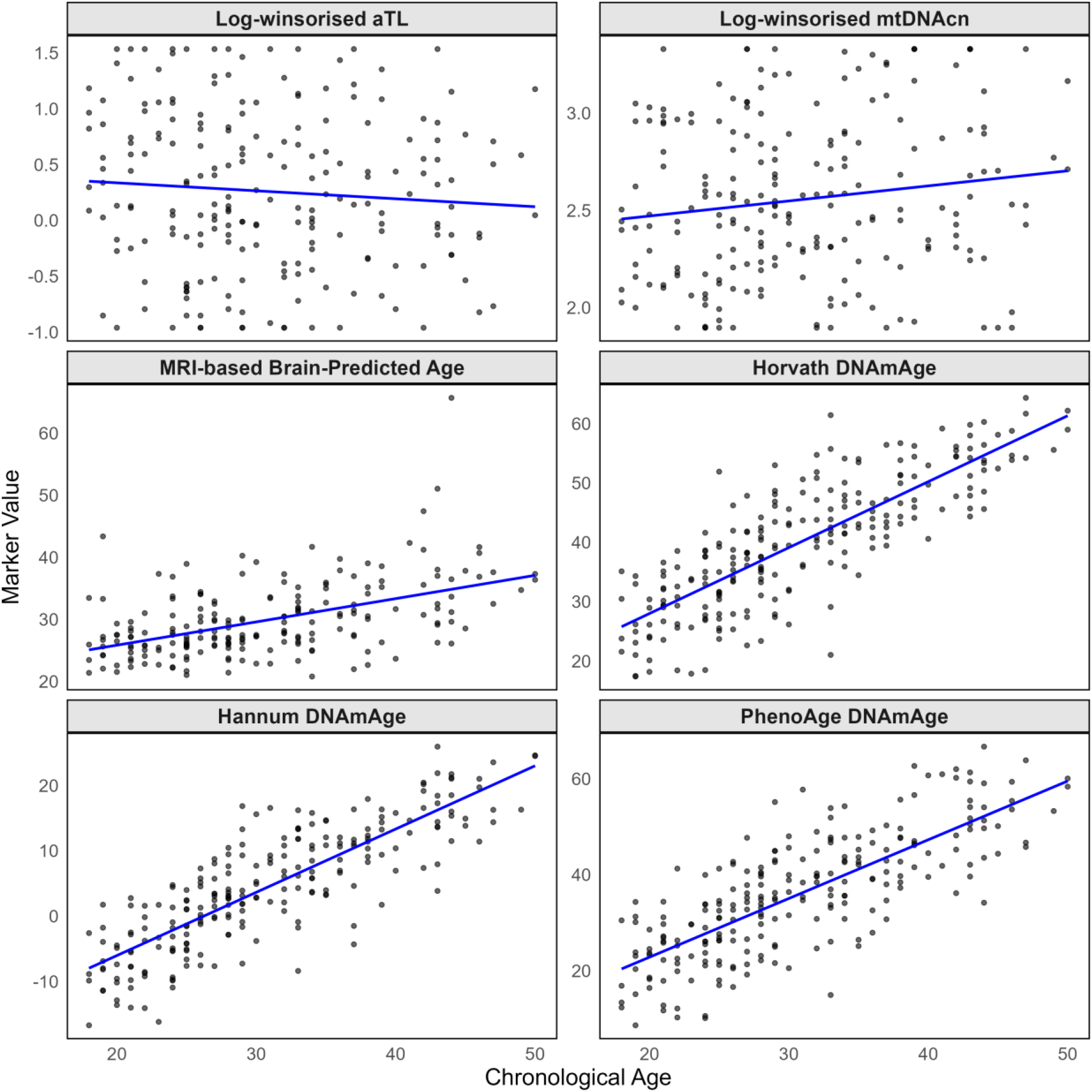
Correlations of biological aging markers against chronological age. Scatterplots show individual data points with fitted linear regression lines. Panels display log-winsorised aTL, log- winsorised mtDNAcn, MRI-BPA, and DNAmAge clocks (Horvath, Hannum, PhenoAge). Log- winsorised aTL showed no significant correlation with age (*r* = –0.08, *p* = 0.249), log-winsorised mtDNAcn was very weakly correlated (*r* = 0.15, *p* = 0.031), MRI-BPA was moderately correlated with chronological age (*r* = 0.50, *p* < 0.001), and DNAmAge clocks were strongly correlated with chronological age (all *r* > 0.77). **Abbreviations:** Absolute telomere length (aTL), DNA methylation age (DNAmAge), magnetic resonance imaging-based brain-predicted age (MRI-BPA), mitochondrial DNA copy number (mtDNAcn)

### Biological aging markers by HIV status

People with HIV demonstrated evidence of accelerated biological aging across several markers (**Figure 2** and **Supplementary Table S2**), including shorter log-winsorised aTL, consistent with the expected age-related telomere shortening (*W* = 7947.50, *r*rb = 0.38, *p* < 0.001), and higher HorvathAA (Welch *t*(225.79) = -4.45, *d* = -0.59, *p* < 0.001), HannumAA (Welch *t*(228.98) = -2.59, *d* = -0.34, *p* = 0.010), PhenoAA (Welch *t*(228.42) = -6.42, *d* = -0.85, *p* < 0.001), and EEAA (Welch *t*(225.70) = -5.80, *d* = -0.76, *p* < 0.001) compared to controls. Unexpectedly, controls showed higher MRI-BPAD values than PWH (Welch *t*(199.90) = 2.54, *d* = 0.36, *p* = 0.012). In contrast, mtDNAcn did not differ significantly between groups.

**Figure 2.**
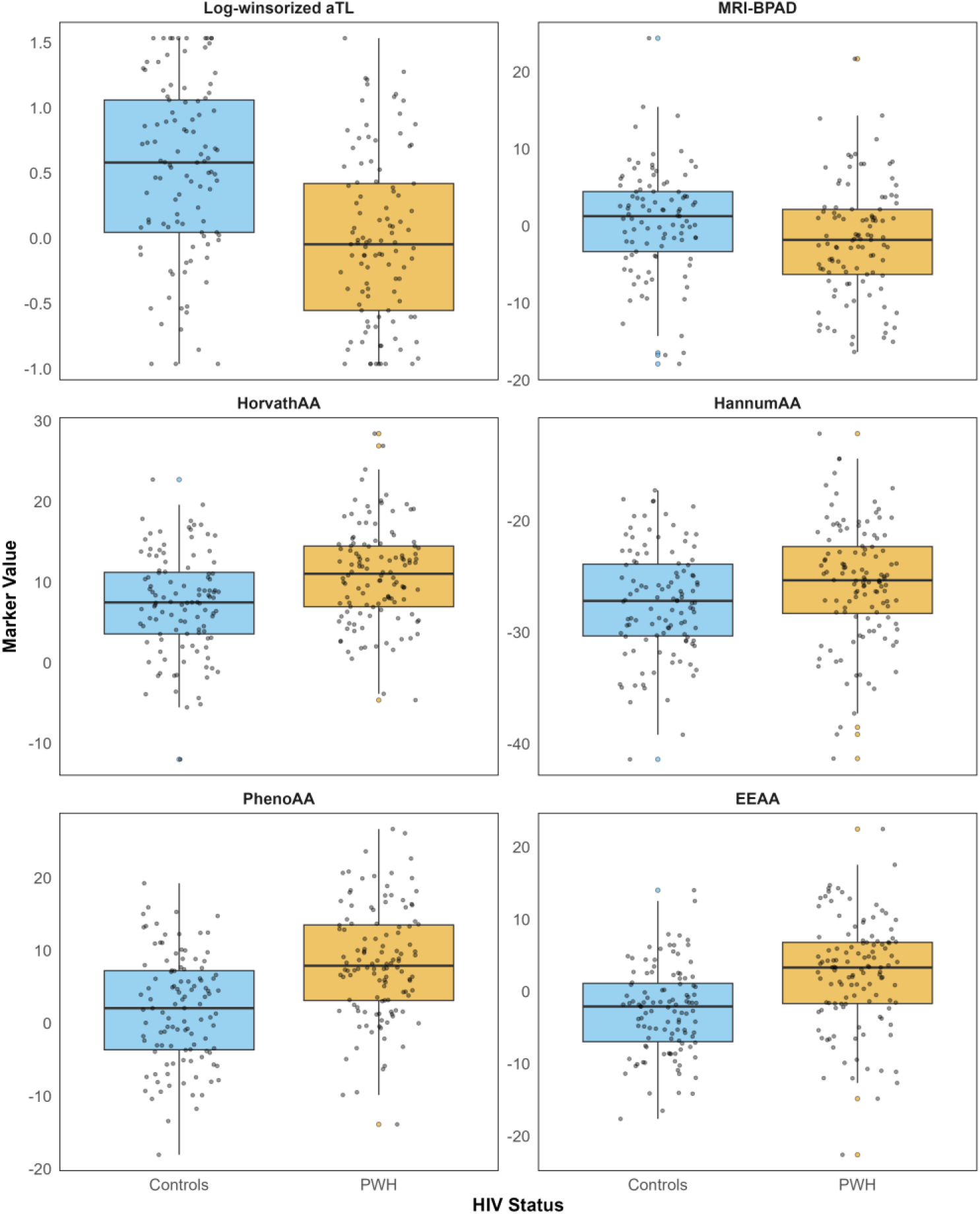
Box plots of aging markers by HIV status (PWH in orange and controls in blue). Log- winsorised aTL (*W* = 7947.50, *rrb* = 0.38, *p* < 0.001), MRI-BPAD (Welch *t*(199.90) = 2.54, *d* = 0.36, *p* = 0.012) were significantly lower in PWH than controls, while HorvathAA (Welch *t*(225.79) = -4.45, *d* = -0.59, *p* < 0.001), HannumAA (Welch *t*(228.98) = -2.59, *d* = -0.34, *p* = 0.010), PhenoAA (Welch *t*(228.42) = -6.42, *d* = -0.85, *p* < 0.001), and EEAA (Welch *t*(225.70) = -5.80, *d* = -0.76, *p* < 0.001) were significantly higher in PWH than controls. mtDNAcn did not differ significantly between groups. **Abbreviations:** Absolute telomere length (aTL), age acceleration (AA), extrinsic epigenetic age acceleration (EEAA), magnetic resonance imaging-based brain-predicted age difference (MRI-BPAD), people with HIV (PWH)

### Biological aging markers and CM scores

Partial Spearman correlations revealed limited evidence of associations between CTQ-SF scores and biological aging markers (**Table 2**). Log-winsorised aTL showed nominal weak correlation with CTQ-SF scores (*ρ* = -0.17, *p* = 0.014), but this did not survive FDR correction. No other markers were significantly associated with CTQ-SF scores.

**Table 2.** Partial Spearman correlation assessing the relationships between aging markers and CTQ-SF scores.

| | $\rho$ | $p$-value | FDR-adjusted $p$-value |
| --- | --- | --- | --- |
| Log-winsorised aTL | -0.17 | 0.014* | 0.130 |
| Log-winsorised mtDNAcn | -0.02 | 0.779 | 0.834 |
| MRI-BPAD | -0.03 | 0.646 | 0.834 |
| HorvathAA | 0.02 | 0.734 | 0.834 |
| HannumAA | -0.04 | 0.514 | 0.834 |
| PhenoAA | 0.07 | 0.302 | 0.834 |
| Horvath IEAA | -0.02 | 0.734 | 0.834 |
| Hannum IEAA | -0.01 | 0.834 | 0.834 |
| EEAA | -0.04 | 0.581 | 0.834 |

**Abbreviations:** Absolute telomere length (aTL), age acceleration (AA), extrinsic epigenetic age acceleration (EEAA), intrinsic epigenetic age acceleration (IEAA), magnetic resonance imaging-based brain-predicted age difference (MRI-BPAD), mitochondrial DNA copy number (mtDNAcn)
**Significance:** \* $p < 0.05$ , \*\* $p < 0.01$ , \*\*\* $p < 0.001$

### Correlations between biological aging markers

Log-winsorised aTL was weakly correlated with log-winsorised mtDNAcn (*ρ* = 0.36, *p* < 0.001) and very weakly correlated with PhenoAA (*ρ* = -0.18, *p* = 0.022) and EEAA (*ρ* = -0.18, *p* = 0.018). MRI-BPAD was weakly correlated with PhenoAA (*r* = 0.20, *p* = 0.012). Moderate to strong intercorrelations were observed among DNAmAge measures, with all pairwise associations statistically significant (*p* < 0.001). The strongest correlations were observed between HorvathAA and Horvath IEAA (*r* = 0.91) and between HannumAA and EEAA (*r* = 0.93). Other DNAmAge measures showed moderate to strong correlations, ranging from *r* = 0.35 for Horvath IEAA and EEAA to *r* = 0.84 for HannumAA and Hannum IEAA (**Figure 3**). Unadjusted *p-*values are provided in **Supplementary Figure S3A** and correlations stratified by HIV status are provided in **Supplementary Figure S3B-C**. Formal tests of whether these associations differed by HIV status are presented in the moderation analyses below.

**Figure 3.**
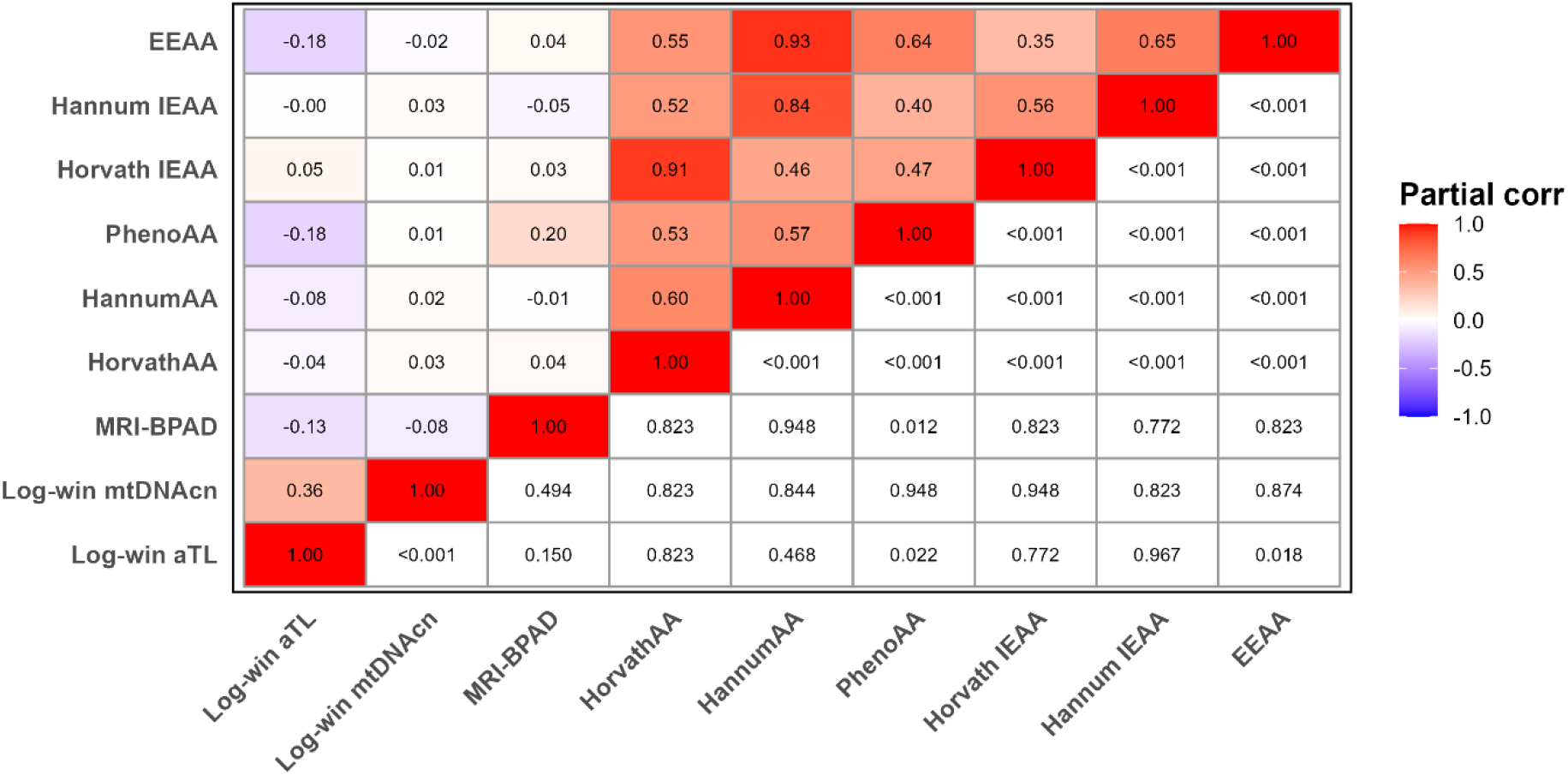
Heatmap of associations among biological aging markers in the overall sample. The upper triangle displays partial correlation coefficients (Pearson or Spearman, depending on marker distribution) with colour intensity reflecting correlation strength and hue indicating direction (blue = negative correlation, red = positive correlation, white = no correlation). The lower triangle displays FDR-adjusted *p*-values. Diagonal cells represent self-correlation (*r*/*ρ* = 1.00). **Abbreviations:** Absolute telomere length (aTL), age acceleration (AA), extrinsic epigenetic age acceleration (EEAA), intrinsic epigenetic age acceleration (IEAA), log-winsorised (Log-win), magnetic resonance imaging-based brain-predicted age difference (MRI-BPAD), mitochondrial DNA copy number (mtDNAcn)

### HIV status and CM scores did not robustly moderate the associations between biological aging markers

Moderation analyses revealed no robust interaction effects after FDR adjustment (**Table 3**). Several nominal associations emerged prior to correction. MRI-BPAD was predicted by the HIV x mtDNAcn interaction (*β* = 12.41, *p* = 0.033), while the HIV x CTQ-SF x mtDNAcn interaction was negatively associated with MRI-BPAD (*β* = -0.29, *p* = 0.021), and the HIV x CTQ-SF x HannumAA was positively associated (*β* = 0.02, *p* = 0.046).

**Table 3.**
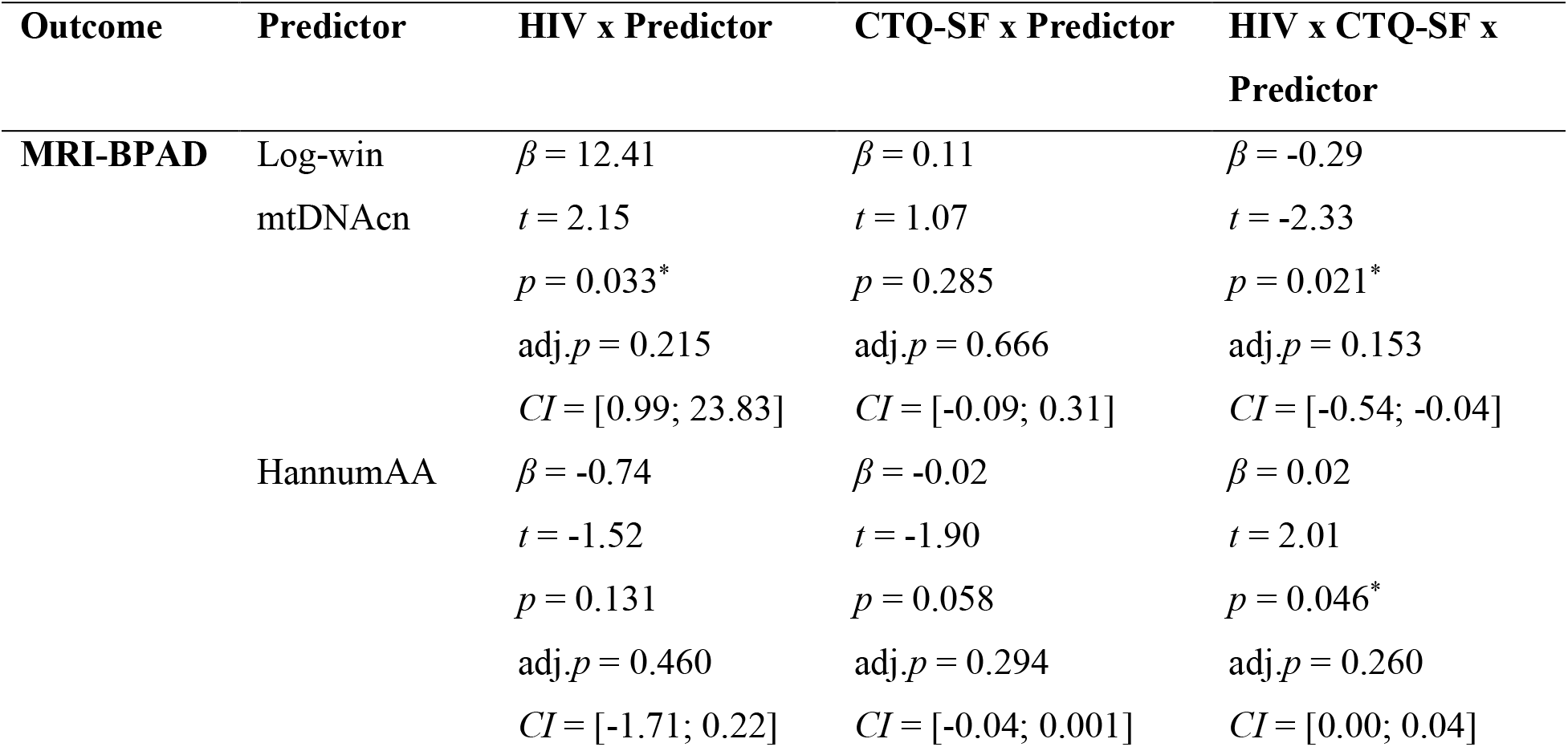
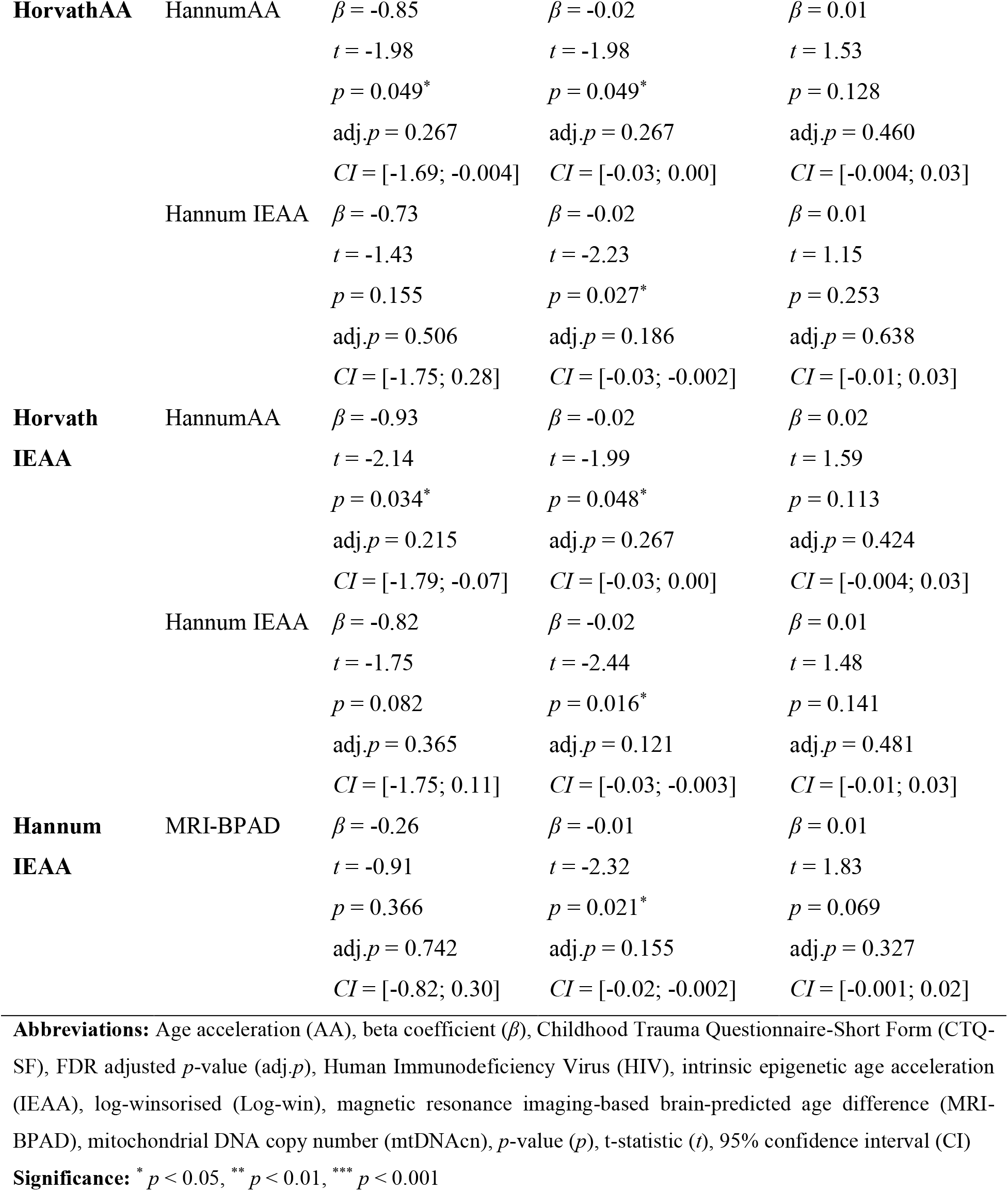
Moderation models testing HIV status and childhood maltreatment effects on associations between biological aging markers.

For HorvathAA, nominal moderation effects were observed for HIV x HannumAA (*β* = -0.85, *p* = 0.049), CTQ-SF x HannumAA (*β* = -0.02, *p* = 0.049), and CTQ-SF x Hannum IEAA (*β* = -0.02, *p* = 0.027). Horvath IEAA showed nominal associations with HIV x HannumAA (*β* = - 0.93, *p* = 0.034), CTQ-SF x HannumAA (*β* = -0.02, *p* = 0.048), and CTQ-SF x Hannum IEAA (*β* = -0.02, *p* = 0.016). Hannum IEAA was nominally associated with CTQ-SF x MRI-BPAD (*β* = -0.01, *p* = 0.021). The full moderation results are provided in **Supplementary Table S3**.

## Discussion

This study examined relationships among nine biological aging markers, capturing molecular, cellular, and neurostructural processes, and tested whether HIV status, CM severity, or their interaction moderated these relationships. Associations across biological aging markers were generally weak, with DNAmAge clocks converging most strongly.

HIV was associated with shorter aTL, unexpectedly lower MRI-BPAD, and accelerated epigenetic aging across multiple DNAmAge clocks (HorvathAA, HannumAA, PhenoAA, and EEAA), while CM did not demonstrate similar effects. These findings are consistent with prior evidence that HIV influences biological aging through telomere attrition, epigenetic alterations, immune dysregulation, and mortality-related pathways (Bożejko et al., 2025; Shiau, Cantos, et al., 2021; Tunnicliffe et al., 2025; Woods et al., 2023). Although moderation analyses did not reveal robust interactions after correction for multiple comparisons, several nominal effects emerged, including HIV x HannumAA and CTQ-SF x HannumAA/IEAA, which were linked to lower HorvathAA. These nominal findings point to possible but inconsistent interactions between HIV, CM, and epigenetic aging that require replication in larger and longitudinal cohorts.

The levels of concordance among epigenetic clocks are consistent with prior evidence that these markers capture overlapping methylation signatures and shared biological aging pathways (Drewelies et al., 2025; Pearce et al., 2022). Importantly, these clocks differ in their training targets and sensitivity to blood-cell composition. The Horvath and Hannum clocks primarily estimate chronological age (Hannum et al., 2013; Horvath, 2013), whereas PhenoAge captures morbidity- and mortality-related physiological aging (Levine et al., 2018). IEAA measures are designed to be relatively independent of inter-individual differences in blood-cell composition, whereas EEAA deliberately incorporates age-related variation in immune-cell composition (e.g., naïve and memory CD8+ T cells, plasmablasts), making them more sensitive to healthspan and immune-related processes (Chen et al., 2016). The overlap we found likely reflects shared biological pathways, particularly those related to immune regulation and inflammation, and is consistent with prior work in people without HIV showing strong clustering within epigenetic markers (Belsky et al., 2017; Kim et al., 2017; Li et al., 2020; Vetter et al., 2019). Future work should test whether this overlap represents a unified methylation aging axis and whether it improves prediction of healthspan and disease risk.

Absolute TL was positively correlated with mtDNAcn, consistent with prior cross-sectional studies reporting co-regulation of these markers across diverse cohorts (Alegría-Torres et al., 2016; Kang et al., 2021; Li et al., 2023; Tyrka et al., 2015). This coupling supports mechanistic links between telomere integrity and mitochondrial function, potentially mediated by p53-dependent signalling and telomerase activity under oxidative stress (Assalve et al., 2025). Evidence from longitudinal work points to reciprocal changes in these systems across time (Hsieh et al., 2020), underscoring the need to interpret cross-sectional correlations as snapshots of complex, evolving cellular processes.

Consistent with previous studies, aTL showed minimal inverse relationships with PhenoAA and EEAA (Chen et al., 2017; Pearce et al., 2022), indicating that shorter telomeres were only weakly associated with greater phenotypic and immune-related epigenetic aging. Immune cell proportions incorporated into EEAA have been linked to telomere dynamics (Chen et al., 2016, 2017), supporting the view that telomere attrition may partly track immune senescence and cumulative biological burden. Taken together, these findings suggest that aTL provides a limited window into the broader biology captured by phenotypic- and immune-related epigenetic clocks, while the weak correlations underscore that these markers are complementary rather than interchangeable.

We found that MRI-BPAD had a limited positive correlation with PhenoAA, our measure of phenotypic biological age acceleration, suggesting a modest overlap between neurostructural aging and phenotypic DNAm signatures. Prior studies have generally reported weak or absent associations between brain age and PhenoAA (Drewelies et al., 2025; Teeuw et al., 2021; Zheng et al., 2021). In contrast, Sant’Anna Barbosa Ferreira et al. (2025) found stronger alignment between HannumAA and MRI-BPAD using the same brain aging model. This difference may reflect cohort composition, as Sant’Anna Barbosa Ferreira et al. (2025) examined a larger, mixed-sex sample of European ancestry. In such populations, HannumAA may track normative neurostructural decline driven by age-related processes such as vascular changes, metabolic stress, and low-grade inflammation (Bektas et al., 2018; Lee & Kim, 2022; Mészáros et al., 2020). By comparison, our cohort comprised younger South African women with relatively preserved immunological profiles, consistent with an early disease stage where these processes may not yet manifest in the brain (Wright et al., 2015). Future studies should test whether MRI-BPAD–HannumAA convergence reflects shared vascular and inflammatory pathways and whether these relationships evolve longitudinally across different disease contexts.

MRI-BPAD was unexpectedly lower in PWH compared to controls, diverging from reports of accelerated brain aging (Cole et al., 2017; Nguchu et al., 2025). Although one possibility is that our brain age model might require further fine-tuning for this population, we consider this unlikely. The model was validated in our sample and demonstrated performance comparable to other studies (Clausen et al., 2022; Han et al., 2021; Hendrikse et al., 2025), supporting its applicability in this context. Importantly, MRI-BPAD did show relationships with phenotypic measures of biological aging, indicating that the model captured relevant aging processes. The unexpectedly lower MRI-BPAD in PWH compared to controls therefore likely reflects a more complex interplay of cohort characteristics and contextual factors. Our cohort was younger, largely ARV-naïve, and showed relatively preserved immunological profiles, which may reflect an early disease stage before neurostructural effects manifest. In contrast, previous brain age studies in PWH include older cohorts, with most participants on HIV treatment (Cole et al., 2017; Underwood et al., 2018). Calibration issues in brain age models, which like epigenetic clocks, have been trained largely in European and North American datasets, may also limit generalisability to African populations (Cole & Franke, 2017; Gibbs et al., 2025; Meeks et al., 2025). Sex-specific differences in brain aging trajectories (Ravndal et al., 2025; Statsenko et al., 2022) and contextual stressors affecting controls could further contribute. Longitudinal studies are needed to clarify whether these dynamics shift over time and how ARV exposure, viral suppression, and inflammation interact with MRI-BPAD trajectories.

We observed no relationship between CM and biological aging markers in this study, which contrasts with meta-analytic evidence linking early adversity to shorter telomeres (Li et al., 2017) and studies associating CM with accelerated epigenetic aging and brain aging (Zhang et al., 2024; Zhou et al., 2025). However, other large-scale studies and meta-analyses have reported null or even biological age-deceleration effects (Meier et al., 2024; Russell et al., 2026). Differences in CM measurement and sample characteristics likely contribute to these discrepancies. For example, Zhang et al. (2024) and Zhou et al. (2025) applied categorical approaches (binary or threshold-based classifications) in larger samples, whereas our study analysed CTQ-SF scores continuously to capture severity across domains, which may be less sensitive in smaller samples (Machlin et al., 2025). Heterogeneity in adversity type, timing, frequency, and psychiatric comorbidities further complicates comparisons (Marini et al., 2020; Russell et al., 2026). Future work would benefit from standardised approaches that integrate both categorical and continuous dimensions to balance sensitivity with comparability (Machlin et al., 2025; Sanchis et al., 2026).

This study has several limitations. Its cross-sectional design precludes inference about longitudinal trajectories of biological aging. Although mtDNAcn qPCR assays showed high amplification efficiency, results should be interpreted with caution due to potential primer-dimer effects and technical variability. The wide confidence intervals observed in moderation models indicate limited precision for detecting small interaction effects, consistent with the need for larger samples in such analyses (Xu et al., 2024). Lastly, we did not examine moderation by HIV clinical markers such as viral load or CD4 count, which may refine understanding of biological aging dynamics (De Francesco et al., 2019). Despite these limitations, the study has several strengths. It employed a multimarker assessment of biological aging; incorporated aTL, which provides a more direct measure of telomere dynamics (O’Callaghan & Fenech, 2011); and validated the MRI-based brain age model, which performed well compared to prior studies (Clausen et al., 2022; Han et al., 2021; Hendrikse et al., 2025). The focus on South African women, a population disproportionately affected by HIV and CM yet underrepresented in aging research, further enhances the study’s contribution.

## Conclusion

Our findings reveal moderate to strong associations among DNAmAge clocks, but weak and inconsistent correlations across other biological aging markers. HIV was linked to accelerated biological aging across several markers, while CM showed limited associations, and neither stressor robustly moderated interrelationships between biological aging markers. These results support a multidimensional model of biological aging and underscore the need for larger, longitudinal, multimarker studies in vulnerable populations.

## Ethical considerations

The study received ethics approval from the Health Research Ethics Committee (HREC) of Stellenbosch University, Cape Town, South Africa, and is renewed annually (project ethics code: N07/07/153). All participants provided informed consent, and the study was conducted in accordance with the recommendations of the Declaration of Helsinki. Authors adhered to the publishing ethics of Global Mental Health.

## Authors’ contributions

**KMSM:** Investigation, formal analysis, data curation, visualisation, writing-original draft, writing-review & editing; **GS:** supervision, methodology, data collection, writing-review & editing; **SD:** supervision, methodology, software, writing-review & editing; **SMJH:** supervision, methodology, writing-review & editing; **SL:** conceptualisation, writing-review & editing; **SS:** conceptualisation, writing-review & editing; **JSW:** conceptualisation, supervision, methodology, funding acquisition, writing-review & editing.

## Data availability statement

The datasets presented in this article are not readily available due to ethical and legal restrictions. Requests to access the datasets should be directed to JSW. The authors are open to collaborating and sharing data within the limits of ethical review restrictions and data transfer policies of Stellenbosch University.

