## Supplementary Figure 1 for "Biological aging markers in South African women with HIV and variable exposure to childhood maltreatment"

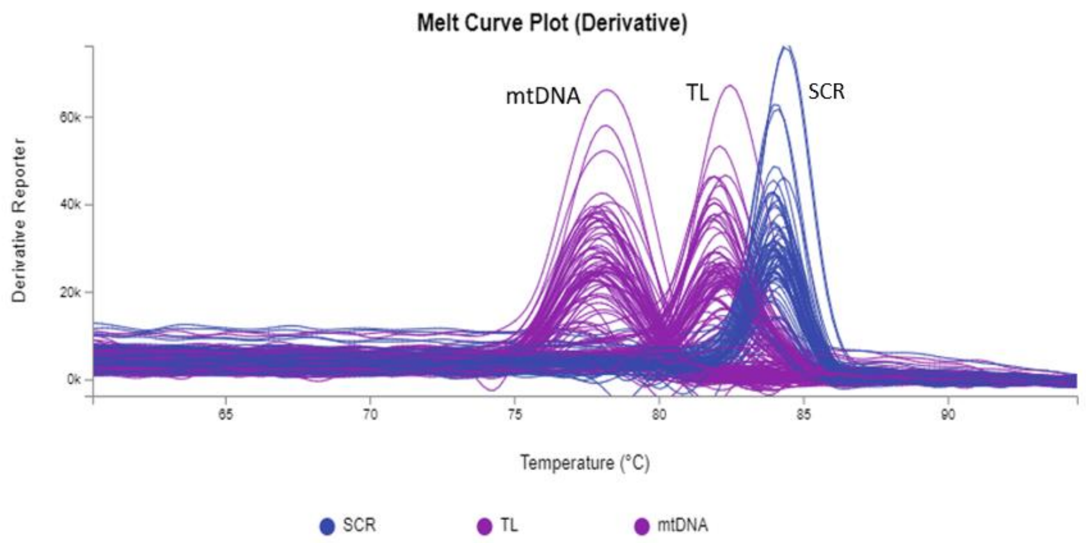


**Figure S1.** Melt curve analysis of qPCR assay. Distinct single peaks were observed for telomere length (TL), mitochondrial DNA copy number (mtDNAcn), and single copy reference (SCR). The derivative plots (–dF/dT vs. temperature) confirm assay specificity and support the reliability of subsequent quantification analyses.
