## Supplementary Figure 2 for "Biological aging markers in South African women with HIV and variable exposure to childhood maltreatment"

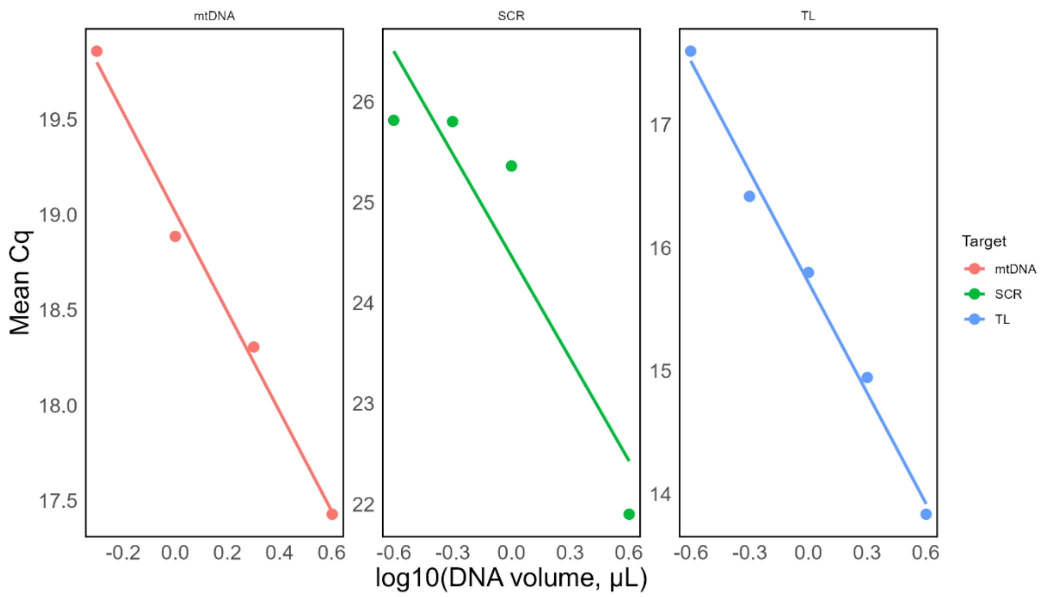


**Figure S2.** Serial dilution of reference DNA. Assay performance metrics are shown for mitochondrial DNA copy number (mtDNAcn; slope = -2.61, $R^2$ = 0.992, efficiency = 141.0%), single copy reference (SCR; slope = -3.38, $R^2$ = 0.846, efficiency = 97.5%), telomere length (TL; slope = -2.99, $R^2$ = 0.991, efficiency = 116.0%).
