## Supplementary Figure 3 for "Biological aging markers in South African women with HIV and variable exposure to childhood maltreatment"

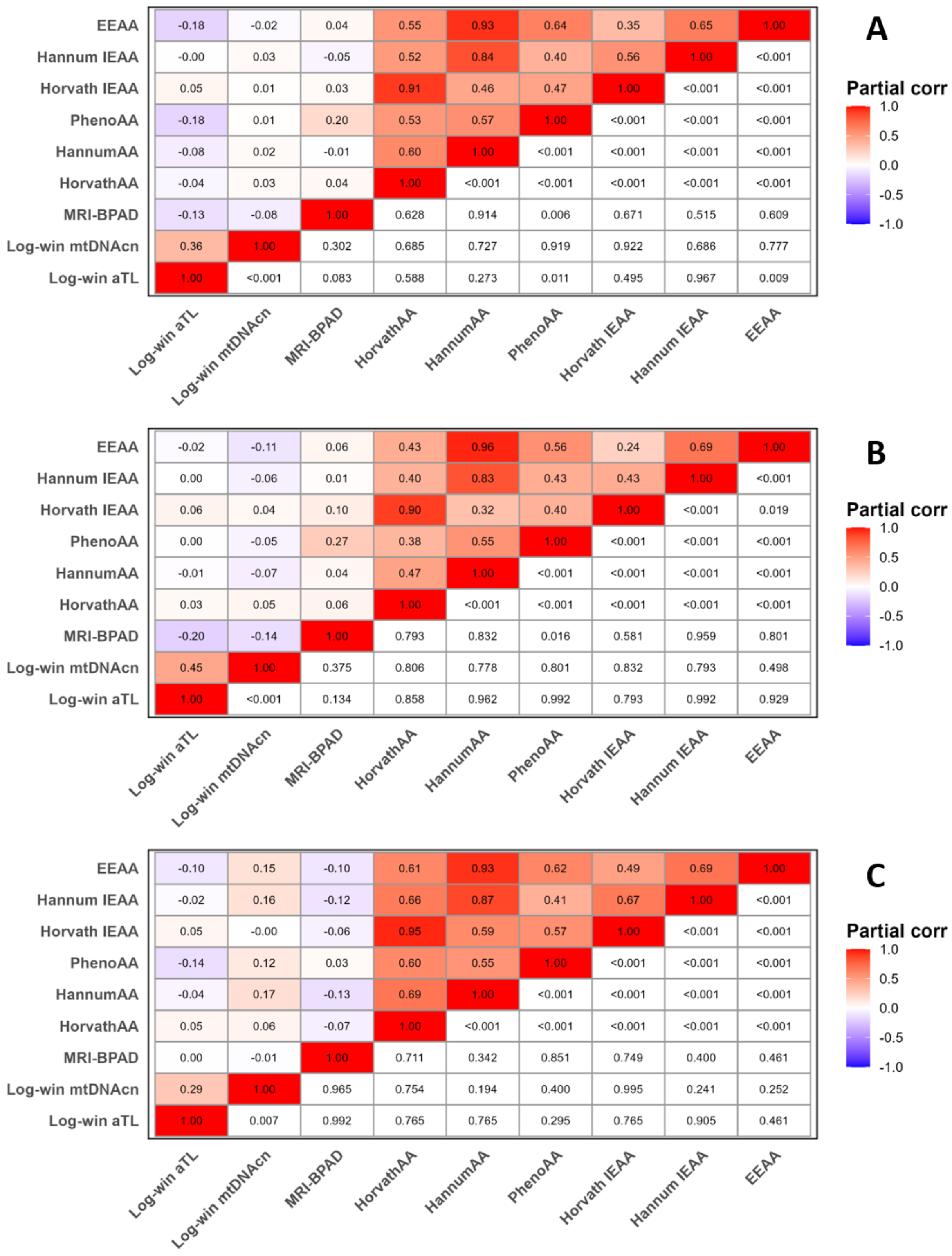


**Figure S3. Heatmaps of associations among biological aging markers in the overall sample and by HIV status.** (**A**) Overall sample, showing partial correlation coefficients and unadjusted *p*-values. (**B**) PWH, showing partial correlation coefficients and FDR-adjusted *p*-values. (**C**) Controls, showing partial correlation coefficients and FDR-adjusted *p*-values. In all panels, the upper triangle displays partial correlation coefficients (Pearson or Spearman, depending on marker distribution), with colour intensity reflecting correlation strength and hue indicating direction (blue = negative correlations, red = positive correlations, white = no correlation). The lower triangle displays *p*-values. Diagonal cells represent self-correlation ($r/\rho$ = 1.00). Partial correlations were adjusted for age, age-squared (MRI-BPAD), MRI protocol type (MRI-BPAD) and plate number (aTL and mtDNAcn). FDR correction was applied separately within each HIV status group. **Abbreviations:** Absolute telomere length (aTL), age acceleration (AA), extrinsic epigenetic age acceleration (EEAA), intrinsic epigenetic age acceleration (IEAA), log-winsorised (Log-win), magnetic resonance imaging-based brain-predicted age difference (MRI-BPAD), mitochondrial DNA copy number (mtDNAcn), people with HIV (PWH).
