## Supplementary Table 1 for "Biological aging markers in South African women with HIV and variable exposure to childhood maltreatment"

**Table S1.** Sample size composition across aging measures

| Measure (s) | Sample size (N) | PWH | Controls |
|---|---|---|---|
| At least 1 measure available | 238 | 122 | 116 |
| aTL + mtDNAcn[1] | 210 | 105 | 105 |
| MRI-BPAD | 202 | 105 | 97 |
| DNAmAge clocks | 231 | 120 | 111 |
| aTL + mtDNAcn + MRI-BPAD | 177 | 90 | 87 |
| aTL + mtDNAcn + DNAmAge clocks | 204 | 104 | 100 |
| MRI-BPAD + DNAmAge clocks | 200 | 104 | 96 |
| aTL + mtDNAcn + MRI-BPAD + DNAmAge clocks | 176 | 90 | 86 |

**Abbreviations:** Absolute telomere length (aTL), age acceleration (AA), extrinsic epigenetic age acceleration (EEAA), intrinsic epigenetic age acceleration (IEAA), magnetic resonance imaging-based brain-predicted age difference (MRI-BPAD), mitochondrial DNA copy number (mtDNAcn).

[1] aTL and mtDNAcn data were available for the same participants (n = 210; 105 PWH, 105 controls)
