## Supplementary Table 2 for "Biological aging markers in South African women with HIV and variable exposure to childhood maltreatment"

**Table S2.** Comparison of biological aging markers between PWH and controls using Welch t-test for normal and Mann-Whitney U test for non-normal markers

| Biological aging measures (years) | Total | | PWH | | Controls | | Statistics |
|---|---|---|---|---|---|---|---|
| | N | mean (SD) | n | mean (SD) | n | mean (SD) | |
| Log-winsorised aTL | 210 | 0.26 (0.71) | 105 | -0.01 (0.64) | 105 | 0.53 (0.67) | $W = 7947.50$, $r_{rb} = 0.38$, $p < 0.001^{***}$ |
| Log-winsorised mtDNAcn | 210 | 2.55 (0.41) | 105 | 2.58 (0.43) | 105 | 2.53 (0.40) | $W = 5140.00$, $r_{rb} = 0.06$, $p = 0.398$ |
| MRI-BPAD | 202 | -0.74 (7.20) | 105 | -1.96 (7.28) | 97 | 0.58 (6.91) | $t(199.86) = 2.54$, $d = 0.36$, $p = 0.012^{*}$ |
| HorvathAA | 231 | 9.18 (6.33) | 120 | 10.90 (5.96) | 111 | 7.33 (6.21) | $t(225.79) = -4.45$, $d = -0.59$, $p < 0.001^{***}$ |
| HannumAA | 231 | -26.40 (5.10) | 120 | -25.57 (5.25) | 111 | -27.29 (4.81) | $t(228.98) = -2.59$, $d = -0.34$, $p = 0.010^{*}$ |
| PhenoAA | 231 | 5.20 (8.20) | 120 | 8.27 (7.66) | 111 | 1.88 (7.45) | $t(228.42) = -6.42$, $d = -0.85$, $p < 0.001^{***}$ |
| Horvath IEAA | 231 | 0.001 (5.69) | 120 | 0.35 (5.52) | 111 | -0.37 (5.86) | $t(224.73) = -0.95$, $d = -0.13$, $p = 0.342$ |
| Hannum IEAA | 231 | 0.006 (4.19) | 120 | 0.17 (4.09) | 111 | -0.17 (4.31) | $t(225.22) = -0.61$, $d = -0.08$, $p = 0.545$ |
| EEAA | 231 | -0.02 (7.20) | 120 | 2.44 (7.36) | 111 | -2.67 (6.02) | $t(225.70) = -5.80$, $d = -0.76$, $p < 0.001^{***}$ |

**Abbreviations:** Absolute telomere length (aTL), age acceleration (AA), Cohen's d (*d*), extrinsic epigenetic age acceleration (EEAA), intrinsic epigenetic age acceleration (IEAA), magnetic resonance imaging-based brain-predicted age difference (MRI-BPAD), Mann-Whitney U test (Wilcoxon rank-sum test) statistic (*W*), mitochondrial DNA copy number (mtDNAcn), rank-biserial correlation ($r_{rb}$), Welch's unequal-variance t-test statistic (*t*).

**Significance:** $^{*}$ $p < 0.05$, $^{**}$ $p < 0.01$, $^{***}$ $p < 0.001$
