## Supplementary Table 3 for "Biological aging markers in South African women with HIV and variable exposure to childhood maltreatment"

**Table S3.** Moderation models testing HIV status and childhood maltreatment effects on associations between biological aging markers

| Outcome | Predictor | HIV x Predictor | CTQ-SF x Predictor | HIV x CTQ-SF x Predictor |
|---|---|---|---|---|
| **Log-win aTL** | Log-win mtDNAcn | $\beta = 0.85$<br>$t = 1.34$<br>$p = 0.183$<br>adj.$p = 0.545$<br>$CI = [-0.41; 2.11]$ | $\beta = 0.001$<br>$t = 0.08$<br>$p = 0.938$<br>adj.$p = 0.986$<br>$CI = [-0.02; 0.02]$ | $\beta = -0.01$<br>$t = -0.76$<br>$p = 0.448$<br>adj.$p = 0.803$<br>$CI = [-0.04; 0.02]$ |
| | MRI-BPAD | $\beta = 0.01$<br>$t = 0.29$<br>$p = 0.776$<br>adj.$p = 0.935$<br>$CI = [-0.08; 0.11]$ | $\beta = 0.00$<br>$t = -0.59$<br>$p = 0.556$<br>adj.$p = 0.877$<br>$CI = [-0.002; 0.001]$ | $\beta = 0.00$<br>$t = -0.35$<br>$p = 0.729$<br>adj.$p = 0.933$<br>$CI = [-0.002; 0.002]$ |
| | HorvathAA | $\beta = 0.03$<br>$t = 0.67$<br>$p = 0.505$<br>adj.$p = 0.839$<br>$CI = [-0.06; 0.12]$ | $\beta = 0.00$<br>$t = 0.27$<br>$p = 0.789$<br>adj.$p = 0.939$<br>$CI = [-0.001; 0.002]$ | $\beta = -0.001$<br>$t = -0.57$<br>$p = 0.572$<br>adj.$p = 0.883$<br>$CI = [-0.002; 0.001]$ |
| | HannumAA | $\beta = 0.09$<br>$t = 1.47$<br>$p = 0.143$<br>adj.$p = 0.484$<br>$CI = [-0.03; 0.21]$ | $\beta = 0.001$<br>$t = 0.89$<br>$p = 0.375$<br>adj.$p = 0.752$<br>$CI = [-0.001; 0.003]$ | $\beta = -0.002$<br>$t = -1.40$<br>$p = 0.164$<br>adj.$p = 0.507$<br>$CI = [-0.004; 0.001]$ |
| | PhenoAA | $\beta = 0.06$<br>$t = 1.60$<br>$p = 0.110$<br>adj.$p = 0.420$<br>$CI = [-0.01; 0.14]$ | $\beta = 0.00$<br>$t = 0.55$<br>$p = 0.585$<br>adj.$p = 0.890$<br>$CI = [-0.001; 0.002]$ | $\beta = -0.001$<br>$t = -1.15$<br>$p = 0.250$<br>adj.$p = 0.637$<br>$CI = [-0.003; 0.001]$ |
| | Horvath IEAA | $\beta = 0.002$<br>$t = 0.04$<br>$p = 0.965$<br>adj.$p = 0.987$<br>$CI = [-0.09; 0.10]$ | $\beta = 0.00$<br>$t = -0.34$<br>$p = 0.733$<br>adj.$p = 0.933$<br>$CI = [-0.002; 0.001]$ | $\beta = 0.00$<br>$t = 0.21$<br>$p = 0.832$<br>adj.$p = 0.953$<br>$CI = [-0.002; 0.002]$ |

| | | | | |
|---|---|---|---|---|
| | Hannum IEAA | *β* = 0.08<br>*t* = 1.17<br>*p* = 0.245<br>adj.*p* = 0.628<br>*CI* = [-0.06; 0.22] | *β* = 0.001<br>*t* = 1.05<br>*p* = 0.297<br>adj.*p* = 0.671<br>*CI* = [-0.001; 0.003] | *β* = -0.002<br>*t* = -1.15<br>*p* = 0.251<br>adj.*p* = 0.637<br>*CI* = [-0.004; 0.001] |
| | EEAA | *β* = 0.07<br>*t* = 1.49<br>*p* = 0.137<br>adj.*p* = 0.474<br>*CI* = [-0.02; 0.17] | *β* = 0.001<br>*t* = 0.53<br>*p* = 0.596<br>adj.*p* = 0.892<br>*CI* = [-0.001; 0.002] | *β* = -0.001<br>*t* = -1.22<br>*p* = 0.226<br>adj.*p* = 0.598<br>*CI* = [-0.003; 0.001] |
| **Log-win mtDNAcn** | Log-win aTL | *β* = 0.26<br>*t* = 1.17<br>*p* = 0.244<br>adj.*p* = 0.628<br>*CI* = [-0.18; 0.70] | *β* = 0.00<br>*t* = 0.07<br>*p* = 0.942<br>adj.*p* = 0.986<br>*CI* = [-0.01; 0.01] | *β* = -0.002<br>*t* = -0.45<br>*p* = 0.654<br>adj.*p* = 0.914<br>*CI* = [-0.01; 0.01] |
| | MRI-BPAD | *β* = 0.03<br>*t* = 1.00<br>*p* = 0.319<br>adj.*p* = 0.699<br>*CI* = [-0.03; 0.09] | *β* = 0.00<br>*t* = 0.32<br>*p* = 0.752<br>adj.*p* = 0.933<br>*CI* = [-0.001; 0.001] | *β* = -0.001<br>*t* = -0.93<br>*p* = 0.353<br>adj.*p* = 0.735<br>*CI* = [-0.002; 0.001] |
| | HorvathAA | *β* = 0.00<br>*t* = -0.01<br>*p* = 0.993<br>adj.*p* = 0.996<br>*CI* = [-0.06; 0.06] | *β* = -0.001<br>*t* = -1.11<br>*p* = 0.267<br>adj.*p* = 0.653<br>*CI* = [-0.002; 0.00] | *β* = 0.00<br>*t* = 0.28<br>*p* = 0.779<br>adj.*p* = 0.935<br>*CI* = [-0.001; 0.001] |
| | HannumAA | *β* = 0.02<br>*t* = 0.67<br>*p* = 0.511<br>adj.*p* = 0.844<br>*CI* = [-0.05; 0.10] | *β* = 0.00<br>*t* = -0.30<br>*p* = 0.763<br>adj.*p* = 0.933<br>*CI* = [-0.001; 0.001] | *β* = -0.001<br>*t* = -0.99<br>*p* = 0.325<br>adj.*p* = 0.707<br>*CI* = [-0.002; 0.001] |
| | PhenoAA | *β* = 0.002<br>*t* = 0.10<br>*p* = 0.919<br>adj.*p* = 0.979<br>*CI* = [-0.05; 0.05] | *β* = -0.001<br>*t* = -1.31<br>*p* = 0.192<br>adj.*p* = 0.555<br>*CI* = [-0.002; 0.00] | *β* = 0.00<br>*t* = 0.004<br>*p* = 0.997<br>adj.*p* = 0.997<br>*CI* = [-0.001; 0.001] |

| | | | | |
|---|---|---|---|---|
| | Horvath IEAA | $\beta = -0.01$ | $\beta = -0.001$ | $\beta = 0.001$ |
| | | $t = -0.41$ | $t = -1.71$ | $t = 0.94$ |
| | | $p = 0.685$ | $p = 0.089$ | $p = 0.350$ |
| | | adj.$p = 0.921$ | adj.$p = 0.381$ | adj.$p = 0.733$ |
| | | $CI = [-0.07; 0.05]$ | $CI = [-0.002; 0.00]$ | $CI = [-0.001; 0.002]$ |
| | Hannum IEAA | $\beta = 0.02$ | $\beta = -0.001$ | $\beta = -0.001$ |
| | | $t = 0.57$ | $t = -0.82$ | $t = -0.70$ |
| | | $p = 0.568$ | $p = 0.414$ | $p = 0.483$ |
| | | adj.$p = 0.883$ | adj.$p = 0.796$ | adj.$p = 0.826$ |
| | | $CI = [-0.06; 0.11]$ | $CI = [-0.002; 0.001]$ | $CI = [-0.002; 0.001]$ |
| | EEAA | $\beta = 0.01$ | $\beta = 0.00$ | $\beta = 0.00$ |
| | | $t = 0.33$ | $t = -0.38$ | $t = -0.60$ |
| | | $p = 0.739$ | $p = 0.707$ | $p = 0.550$ |
| | | adj.$p = 0.933$ | adj.$p = 0.933$ | adj.$p = 0.871$ |
| | | $CI = [-0.05; 0.07]$ | $CI = [-0.001; 0.001]$ | $CI = [-0.002; 0.001]$ |
| **MRI-BPAD** | Log-win aTL | $\beta = 2.68$ | $\beta = 0.01$ | $\beta = -0.09$ |
| | | $t = 0.80$ | $t = 0.10$ | $t = -1.30$ |
| | | $p = 0.422$ | $p = 0.922$ | $p = 0.194$ |
| | | adj.$p = 0.799$ | adj.$p = 0.979$ | adj.$p = 0.560$ |
| | | $CI = [-3.90; 9.25]$ | $CI = [-0.09; 0.10]$ | $CI = [-0.22; 0.04]$ |
| | Log-win mtDNAcn | $\beta = 12.41$ | $\beta = 0.11$ | $\beta = -0.29$ |
| | | $t = 2.15$ | $t = 1.07$ | $t = -2.33$ |
| | | $p = 0.033^{*}$ | $p = 0.285$ | $p = 0.021^{*}$ |
| | | adj.$p = 0.215$ | adj.$p = 0.666$ | adj.$p = 0.153$ |
| | | $CI = [0.99; 23.83]$ | $CI = [-0.09; 0.31]$ | $CI = [-0.54; -0.04]$ |
| | HorvathAA | $\beta = -0.53$ | $\beta = -0.01$ | $\beta = 0.01$ |
| | | $t = -1.41$ | $t = -0.91$ | $t = 1.68$ |
| | | $p = 0.159$ | $p = 0.364$ | $p = 0.095$ |
| | | adj.$p = 0.507$ | adj.$p = 0.742$ | adj.$p = 0.396$ |
| | | $CI = [-1.26; 0.21]$ | $CI = [-0.02; 0.01]$ | $CI = [-0.002; 0.03]$ |
| | HannumAA | $\beta = -0.74$ | $\beta = -0.02$ | $\beta = 0.02$ |
| | | $t = -1.52$ | $t = -1.90$ | $t = 2.01$ |
| | | $p = 0.131$ | $p = 0.058$ | $p = 0.046^{*}$ |
| | | adj.$p = 0.460$ | adj.$p = 0.294$ | adj.$p = 0.260$ |
| | | $CI = [-1.71; 0.22]$ | $CI = [-0.04; 0.001]$ | $CI = [0.00; 0.04]$ |

| | | | | |
|---|---|---|---|---|
| | PhenoAA | $\beta = -0.24$ | $\beta = -0.01$ | $\beta = 0.01$ |
| | | $t = -0.80$ | $t = -0.88$ | $t = 1.39$ |
| | | $p = 0.426$ | $p = 0.378$ | $p = 0.166$ |
| | | adj.$p = 0.799$ | adj.$p = 0.757$ | adj.$p = 0.507$ |
| | | $CI$ = [-0.85; 0.36] | $CI$ = [-0.02; 0.01] | $CI$ = [-0.004; 0.02] |
| | Horvath IEAA | $\beta = -0.50$ | $\beta = -0.01$ | $\beta = 0.01$ |
| | | $t = -1.27$ | $t = -0.70$ | $t = 1.54$ |
| | | $p = 0.206$ | $p = 0.487$ | $p = 0.125$ |
| | | adj.$p = 0.568$ | adj.$p = 0.826$ | adj.$p = 0.457$ |
| | | $CI$ = [-1.28; 0.28] | $CI$ = [-0.02; 0.01] | $CI$ = [-0.004; 0.03] |
| | Hannum IEAA | $\beta = -0.86$ | $\beta = -0.02$ | $\beta = 0.02$ |
| | | $t = -1.52$ | $t = -1.86$ | $t = 1.95$ |
| | | $p = 0.130$ | $p = 0.064$ | $p = 0.053$ |
| | | adj.$p = 0.460$ | adj.$p$ =0.311 | adj.$p = 0.280$ |
| | | $CI$ = [-1.97; 0.26] | $CI$ = [-0.04; 0.001] | $CI$ = [0.00; 0.05] |
| | EEAA | $\beta = -0.57$ | $\beta = -0.02$ | $\beta = 0.02$ |
| | | $t = -1.50$ | $t = -1.93$ | $t = 1.96$ |
| | | $p = 0.136$ | $p = 0.056$ | $p = 0.051$ |
| | | adj.$p = 0.474$ | adj.$p = 0.288$ | adj.$p = 0.275$ |
| | | $CI$ = [-1.33; 0.18] | $CI$ = [-0.03; 0.00] | $CI$ = [0.00; 0.03] |
| **HorvathAA** | Log-win aTL | $\beta = 0.96$ | $\beta = 0.01$ | $\beta = -0.03$ |
| | | $t = 0.25$ | $t = 0.21$ | $t = -0.34$ |
| | | $p = 0.803$ | $p = 0.836$ | $p = 0.735$ |
| | | adj.$p = 0.943$ | adj.$p = 0.953$ | adj.$p = 0.933$ |
| | | $CI$ = [-6.63; 8.55] | $CI$ = [-0.10; 0.13] | $CI$ = [-0.18; 0.13] |
| | Log-win mtDNAcn | $\beta = -2.00$ | $\beta = -0.17$ | $\beta = 0.09$ |
| | | $t = -0.29$ | $t = -1.32$ | $t = 0.602$ |
| | | $p = 0.771$ | $p = 0.190$ | $p = 0.548$ |
| | | adj.$p = 0.935$ | adj.$p = 0.555$ | adj.$p = 0.871$ |
| | | $CI$ = [-15.54; 11.55] | $CI$ = [-0.42; 0.08] | $CI$ = [-0.21; 0.39] |
| | MRI-BPAD | $\beta = -0.09$ | $\beta = -0.003$ | $\beta = 0.01$ |
| | | $t = -0.23$ | $t = -0.54$ | $t = 0.62$ |
| | | $p = 0.820$ | $p = 0.590$ | $p = 0.539$ |
| | | adj.$p = 0.949$ | adj.$p = 0.890$ | adj.$p = 0.867$ |
| | | $CI$ = [-0.90; 0.72] | $CI$ = [-0.02; 0.01] | $CI$ = [-0.01; 0.02] |

| | | | | |
|---|---|---|---|---|
| | HannumAA | $\beta = -0.85$<br>$t = -1.98$<br>$p = 0.049^{*}$<br>adj.$p = 0.267$<br>$CI = [-1.69; -0.004]$ | $\beta = -0.02$<br>$t = -1.98$<br>$p = 0.049^{*}$<br>adj.$p = 0.267$<br>$CI = [-0.03; 0.00]$ | $\beta = 0.01$<br>$t = 1.53$<br>$p = 0.128$<br>adj.$p = 0.460$<br>$CI = [-0.004; 0.03]$ |
| | PhenoAA | $\beta = -0.36$<br>$t = -1.17$<br>$p = 0.242$<br>adj.$p = 0.628$<br>$CI = [-0.96; 0.24]$ | $\beta = -0.001$<br>$t = -0.1$<br>$p = 0.921$<br>adj.$p = 0.979$<br>$CI = [-0.01; 0.01]$ | $\beta = 0.003$<br>$t = 0.41$<br>$p = 0.680$<br>adj.$p = 0.916$<br>$CI = [-0.01; 0.02]$ |
| | Horvath IEAA | $\beta = 0.02$<br>$t = 0.11$<br>$p = 0.913$<br>adj.$p = 0.978$<br>$CI = [-0.31; 0.35]$ | $\beta = -0.001$<br>$t = -0.16$<br>$p = 0.870$<br>adj.$p = 0.969$<br>$CI = [-0.01; 0.01]$ | $\beta = -0.001$<br>$t = -0.21$<br>$p = 0.834$<br>adj.$p = 0.953$<br>$CI = [-0.01; 0.01]$ |
| | Hannum IEAA | $\beta = -0.73$<br>$t = -1.43$<br>$p = 0.155$<br>adj.$p = 0.506$<br>$CI = [-1.75; 0.28]$ | $\beta = -0.02$<br>$t = -2.23$<br>$p = 0.027^{*}$<br>adj.$p = 0.186$<br>$CI = [-0.03; -0.002]$ | $\beta = 0.01$<br>$t = 1.15$<br>$p = 0.253$<br>adj.$p = 0.638$<br>$CI = [-0.01; 0.03]$ |
| | EEAA | $\beta = -0.65$<br>$t = -1.82$<br>$p = 0.070$<br>adj.$p = 0.327$<br>$CI = [-1.36; 0.05]$ | $\beta = -0.01$<br>$t = -1.40$<br>$p = 0.163$<br>adj.$p = 0.507$<br>$CI = [-0.02; 0.004]$ | $\beta = 0.01$<br>$t = 1.25$<br>$p = 0.213$<br>adj.$p = 0.576$<br>$CI = [-0.01; 0.03]$ |
| **HannumAA** | Log-win aTL | $\beta = 3.46$<br>$t = 1.14$<br>$p = 0.254$<br>adj.$p = 0.639$<br>$CI = [-2.51; 9.44]$ | $\beta = 0.05$<br>$t = 1.10$<br>$p = 0.273$<br>adj.$p = 0.656$<br>$CI = [-0.04; 0.14]$ | $\beta = -0.09$<br>$t = -1.46$<br>$p = 0.147$<br>adj.$p = 0.489$<br>$CI = [-0.21; 0.03]$ |
| | Log-win mtDNAcn | $\beta = 2.30$<br>$t = 0.43$<br>$p = 0.669$<br>adj.$p = 0.914$<br>$CI = [-8.31; 12.91]$ | $\beta = -0.04$<br>$t = -0.39$<br>$p = 0.701$<br>adj.$p = 0.930$<br>$CI = [-0.23; 0.16]$ | $\beta = -0.09$<br>$t = -0.73$<br>$p = 0.466$<br>adj.$p = 0.816$<br>$CI = [-0.32; 0.15]$ |

| | | | | |
|---|---|---|---|---|
| | MRI-BPAD | $\beta = -0.33$ | $\beta = -0.01$ | $\beta = 0.01$ |
| | | $t = -0.98$ | $t = -1.53$ | $t = 1.60$ |
| | | $p = 0.331$ | $p = 0.128$ | $p = 0.111$ |
| | | adj.$p = 0.712$ | adj.$p = 0.460$ | adj.$p = 0.422$ |
| | | $CI = [-1.00; 0.34]$ | $CI = [-0.02; 0.002]$ | $CI = [-0.002; 0.02]$ |
| | HorvathAA | $\beta = -0.37$ | $\beta = -0.004$ | $\beta = 0.01$ |
| | | $t = -1.35$ | $t = -0.80$ | $t = 0.89$ |
| | | $p = 0.177$ | $p = 0.426$ | $p = 0.374$ |
| | | adj.$p = 0.530$ | adj.$p = 0.799$ | adj.$p = 0.751$ |
| | | $CI = [-0.90; 0.17]$ | $CI = [-0.01; 0.01]$ | $CI = [-0.01; 0.02]$ |
| | PhenoAA | $\beta = -0.17$ | $\beta = -0.01$ | $\beta = 0.004$ |
| | | $t = -0.71$ | $t = -0.97$ | $t = 0.83$ |
| | | $p = 0.481$ | $p = 0.334$ | $p = 0.406$ |
| | | adj.$p = 0.826$ | adj.$p = 0.714$ | adj.$p = 0.792$ |
| | | $CI = [-0.64; 0.31]$ | $CI = [-0.01; 0.01]$ | $CI = [-0.01; 0.02]$ |
| | Horvath IEAA | $\beta = -0.47$ | $\beta = -0.01$ | $\beta = 0.01$ |
| | | $t = -1.47$ | $t = -1.12$ | $t = 1.02$ |
| | | $p = 0.144$ | $p = 0.265$ | $p = 0.308$ |
| | | adj.$p = 0.484$ | adj.$p = 0.651$ | adj.$p = 0.680$ |
| | | $CI = [-1.10; 0.16]$ | $CI = [-0.02; 0.01]$ | $CI = [-0.01; 0.02]$ |
| | Hannum IEAA | $\beta = 0.28$ | $\beta = -0.002$ | $\beta = -0.003$ |
| | | $t = 1.06$ | $t = -0.52$ | $t = -0.63$ |
| | | $p = 0.291$ | $p = 0.605$ | $p = 0.533$ |
| | | adj.$p = 0.669$ | adj.$p = 0.898$ | adj.$p = 0.863$ |
| | | $CI = [-0.24; 0.80]$ | $CI = [-0.01; 0.01]$ | $CI = [-0.01; 0.01]$ |
| | EEAA | $\beta = -0.07$ | $\beta = 0.00$ | $\beta = 0.00$ |
| | | $t = -0.67$ | $t = 0.01$ | $t = 0.07$ |
| | | $p = 0.504$ | $p = 0.988$ | $p = 0.943$ |
| | | adj.$p = 0.839$ | adj.$p = 0.992$ | adj.$p = 0.986$ |
| | | $CI = [-0.29; 0.14]$ | $CI = [-0.004; 0.004]$ | $CI = [-0.01; 0.01]$ |
| **PhenoAA** | Log-win aTL | $\beta = 6.53$ | $\beta = 0.04$ | $\beta = -0.12$ |
| | | $t = 1.40$ | $t = 0.54$ | $t = -1.29$ |
| | | $p = 0.164$ | $p = 0.591$ | $p = 0.198$ |
| | | adj.$p = 0.507$ | adj.$p = 0.890$ | adj.$p = 0.564$ |
| | | $CI = [-2.70; 15.76]$ | $CI = [-0.10; 0.18]$ | $CI = [-0.31; 0.06]$ |

| | | | |
|---|---|---|---|
| Log-win | $\beta = 2.97$ | $\beta = -0.21$ | $\beta = -0.03$ |
| mtDNAcn | $t = 0.36$ | $t = -1.40$ | $t = -0.16$ |
| | $p = 0.720$ | $p = 0.163$ | $p = 0.874$ |
| | adj.$p = 0.933$ | adj.$p = 0.507$ | adj.$p = 0.972$ |
| | $CI = [-13.35; 19.30]$ | $CI = [-0.51; 0.09]$ | $CI = [-0.38; 0.33]$ |
| MRI-BPAD | $\beta = -0.20$ | $\beta = -0.003$ | $\beta = 0.01$ |
| | $t = -0.40$ | $t = -0.42$ | $t = 0.75$ |
| | $p = 0.691$ | $p = 0.678$ | $p = 0.457$ |
| | adj.$p = 0.922$ | adj.$p = 0.916$ | adj.$p = 0.811$ |
| | $CI = [-1.21; 0.80]$ | $CI = [-0.02; 0.01]$ | $CI = [-0.01; 0.03]$ |
| HorvathAA | $\beta = -0.52$ | $\beta = 0.01$ | $\beta = 0.003$ |
| | $t = -1.19$ | $t = 0.82$ | $t = 0.36$ |
| | $p = 0.237$ | $p = 0.415$ | $p = 0.722$ |
| | adj.$p = 0.623$ | adj.$p = 0.796$ | adj.$p = 0.933$ |
| | $CI = [-1.38; 0.34]$ | $CI = [-0.01; 0.02]$ | $CI = [-0.02; 0.02]$ |
| HannumAA | $\beta = -0.59$ | $\beta = -0.01$ | $\beta = 0.01$ |
| | $t = -1.08$ | $t = -0.68$ | $t = 1.09$ |
| | $p = 0.283$ | $p = 0.495$ | $p = 0.278$ |
| | adj.$p = 0.664$ | adj.$p = 0.832$ | adj.$p = 0.660$ |
| | $CI = [-1.68; 0.49]$ | $CI = [-0.03; 0.01]$ | $CI = [-0.01; 0.04]$ |
| Horvath IEAA | $\beta = -0.35$ | $\beta = 0.01$ | $\beta = 0.001$ |
| | $t = -0.74$ | $t = 0.87$ | $t = 0.11$ |
| | $p = 0.462$ | $p = 0.385$ | $p = 0.909$ |
| | adj.$p = 0.813$ | adj.$p = 0.766$ | adj.$p = 0.978$ |
| | $CI = [-1.30; 0.59]$ | $CI = [-0.01; 0.03]$ | $CI = [-0.02; 0.02]$ |
| Hannum IEAA | $\beta = -0.33$ | $\beta = -0.01$ | $\beta = 0.01$ |
| | $t = -0.48$ | $t = -1.17$ | $t = 0.82$ |
| | $p = 0.634$ | $p = 0.245$ | $p = 0.411$ |
| | adj.$p = 0.914$ | adj.$p = 0.628$ | adj.$p = 0.795$ |
| | $CI = [-1.70; 1.04]$ | $CI = [-0.03; 0.01]$ | $CI = [-0.02; 0.04]$ |
| EEAA | $\beta = -0.33$ | $\beta = 0.002$ | $\beta = 0.003$ |
| | $t = -0.79$ | $t = 0.20$ | $t = 0.31$ |
| | $p = 0.433$ | $p = 0.843$ | $p = 0.755$ |
| | adj.$p = 0.801$ | adj.$p = 0.953$ | adj.$p = 0.933$ |
| | $CI = [-1.16; 0.50]$ | $CI = [-0.02; 0.02]$ | $CI = [-0.02; 0.02]$ |

| | | | | |
|---|---|---|---|---|
| **Horvath IEAA** | Log-win aTL | *β* = -1.24<br>*t* = -0.34<br>*p* = 0.732<br>adj.*p* = 0.933<br>*CI* = [-8.38; 5.90] | *β* = -0.01<br>*t* = -0.18<br>*p* = 0.859<br>adj.*p* = 0.960<br>*CI* = [-0.12; 0.10] | *β* = 0.03<br>*t* = 0.39<br>*p* = 0.700<br>adj.*p* =0.930<br>*CI* = [-0.12; 0.17] |
| | Log-win mtDNAcn | *β* = -2.94<br>*t* = -0.46<br>*p* = 0.649<br>adj.*p* = 0.914<br>*CI* = [-15.62; 9.75] | *β* = -0.20<br>*t* = -1.71<br>*p* = 0.090<br>adj.*p* = 0.381<br>*CI* = [-0.44; 0.03] | *β* = 0.13<br>*t* = 0.94<br>*p* = 0.347<br>adj.*p* = 0.732<br>*CI* = [-0.14; 0.41] |
| | MRI-BPAD | *β* = 0.15<br>*t* = 0.38<br>*p* = 0.703<br>adj.*p* = 0.930<br>*CI* = [-0.62; 0.92] | *β* = -0.002<br>*t* = -0.28<br>*p* = 0.779<br>adj.*p* = 0.935<br>*CI* = [-0.01; 0.01] | *β* = 0.001<br>*t* = 0.177<br>*p* = 0.859<br>adj.*p* = 0.960<br>*CI* = [-0.01; 0.02] |
| | HorvathAA | *β* = -0.12<br>*t* = -0.83<br>*p* = 0.410<br>adj.*p* = 0.795<br>*CI* = [-0.40; 0.17] | *β* = -0.001<br>*t* = -0.31<br>*p* = 0.759<br>adj.*p* = 0.933<br>*CI* = [-0.01; 0.004] | *β* = 0.001<br>*t* = 0.29<br>*p* = 0.776<br>adj.*p* = 0.935<br>*CI* = [-0.01; 0.01] |
| | HannumAA | *β* = -0.93<br>*t* = -2.14<br>*p* = 0.034*<br>adj.*p* = 0.215<br>*CI* = [-1.79; -0.07] | *β* = -0.02<br>*t* = -1.99<br>*p* = 0.048*<br>adj.*p* = 0.267<br>*CI* = [-0.03; 0.00] | *β* = 0.02<br>*t* = 1.59<br>*p* = 0.113<br>adj.*p* = 0.424<br>*CI* = [-0.004; 0.03] |
| | PhenoAA | *β* = -0.32<br>*t* = -1.10<br>*p* = 0.273<br>adj.*p* = 0.656<br>*CI* = [-0.88; 0.25] | *β* = -0.001<br>*t* = -0.14<br>*p* = 0.890<br>adj.*p* = 0.978<br>*CI* = [-0.01; 0.01] | *β* = 0.003<br>*t* = 0.44<br>*p* = 0.660<br>adj.*p* = 0.914<br>*CI* = [-0.01; 0.02] |
| | Hannum IEAA | *β* = -0.82<br>*t* = -1.75<br>*p* = 0.082<br>adj.*p* = 0.365<br>*CI* = [-1.75; 0.11] | *β* = -0.02<br>*t* = -2.44<br>*p* = 0.016*<br>adj.*p* = 0.121<br>*CI* = [-0.03; -0.003] | *β* = 0.01<br>*t* = 1.48<br>*p* = 0.141<br>adj.*p* = 0.481<br>*CI* = [-0.01; 0.03] |

| | | | | |
|---|---|---|---|---|
| | EEAA | $\beta = -0.70$<br>$t = -1.91$<br>$p = 0.057$<br>adj.$p = 0.293$<br>$CI = [-1.41; 0.02]$ | $\beta = -0.01$<br>$t = -1.41$<br>$p = 0.160$<br>adj.$p = 0.507$<br>$CI = [-0.03; 0.004]$ | $\beta = 0.01$<br>$t = 1.28$<br>$p = 0.201$<br>adj.$p = 0.564$<br>$CI = [-0.01; 0.03]$ |
| **Hannum IEAA** | Log-win aTL | $\beta = 1.42$<br>$t = 0.55$<br>$p = 0.584$<br>adj.$p = 0.890$<br>$CI = [-3.70; 6.55]$ | $\beta = 0.04$<br>$t = 1.10$<br>$p = 0.273$<br>adj.$p = 0.656$<br>$CI = [-0.03; 0.12]$ | $\beta = -0.04$<br>$t = -0.78$<br>$p = 0.439$<br>adj.$p = 0.801$<br>$CI = [-0.14; 0.06]$ |
| | Log-win mtDNAcn | $\beta = 1.30$<br>$t = 0.28$<br>$p = 0.778$<br>adj.$p = 0.935$<br>$CI = [-7.74; 10.33]$ | $\beta = -0.07$<br>$t = -0.80$<br>$p = 0.424$<br>adj.$p = 0.935$<br>$CI = [-0.23; 0.10]$ | $\beta = -0.05$<br>$t = -0.49$<br>$p = 0.626$<br>adj.$p = 0.911$<br>$CI = [-0.25; 0.15]$ |
| | MRI-BPAD | $\beta = -0.26$<br>$t = -0.91$<br>$p = 0.366$<br>adj.$p = 0.742$<br>$CI = [-0.82; 0.30]$ | $\beta = -0.01$<br>$t = -2.32$<br>$p = 0.021^{*}$<br>adj.$p = 0.155$<br>$CI = [-0.02; -0.002]$ | $\beta = 0.01$<br>$t = 1.83$<br>$p = 0.069$<br>adj.$p = 0.327$<br>$CI = [-0.001; 0.02]$ |
| | HorvathAA | $\beta = -0.33$<br>$t = -1.40$<br>$p = 0.163$<br>adj.$p = 0.507$<br>$CI = [-0.80; 0.14]$ | $\beta = -0.003$<br>$t = -0.71$<br>$p = 0.479$<br>adj.$p = 0.826$<br>$CI = [-0.01; 0.01]$ | $\beta = 0.003$<br>$t = 0.65$<br>$p = 0.518$<br>adj.$p = 0.850$<br>$CI = [-0.01; 0.01]$ |
| | HannumAA | $\beta = -0.10$<br>$t = -0.50$<br>$p = 0.619$<br>adj.$p = 0.909$<br>$CI = [-0.48; 0.28]$ | $\beta = 0.001$<br>$t = 0.35$<br>$p = 0.724$<br>adj.$p = 0.933$<br>$CI = [-0.01; 0.01]$ | $\beta = 0.00$<br>$t = -0.11$<br>$p = 0.916$<br>adj.$p = 0.978$<br>$CI = [-0.01; 0.01]$ |
| | PhenoAA | $\beta = -0.17$<br>$t = -0.79$<br>$p = 0.433$<br>adj.$p = 0.801$<br>$CI = [-0.61; 0.26]$ | $\beta = -0.004$<br>$t = -0.98$<br>$p = 0.330$<br>adj.$p = 0.712$<br>$CI = [-0.01; 0.004]$ | $\beta = 0.004$<br>$t = 0.80$<br>$p = 0.426$<br>adj.$p = 0.799$<br>$CI = [-0.01; 0.01]$ |

| | | | | |
|---|---|---|---|---|
| | Horvath IEAA | *β* = -0.39<br>*t* = -1.54<br>*p* = 0.126<br>adj.*p* = 0.459<br>*CI* = [-0.88; 0.11] | *β* = -0.01<br>*t* = -1.03<br>*p* = 0.303<br>adj.*p* = 0.677<br>*CI* = [-0.01; 0.004] | *β* = 0.01<br>*t* = 1.00<br>*p* = 0.319<br>adj.*p* = 0.699<br>*CI* = [-0.01; 0.02] |
| | EEAA | *β* = -0.10<br>*t* = -0.49<br>*p* = 0.624<br>adj.*p* = 0.910<br>*CI* = [-0.51; 0.31] | *β* = 0.00<br>*t* = -0.09<br>*p* = 0.930<br>adj.*p* = 0.983<br>*CI* = [-0.01; 0.01] | *β* = 0.00<br>*t* = 0.05<br>*p* = 0.958<br>adj.*p* = 0.987<br>*CI* = [-0.01; 0.01] |
| **EEAA** | Log-win aTL | *β* = 5.57<br>*t* = 1.40<br>*p* = 0.162<br>adj.*p* = 0.507<br>*CI* = [-2.26; 13.40] | *β* = 0.05<br>*t* = 0.78<br>*p* = 0.437<br>adj.*p* = 0.801<br>*CI* = [-0.07; 0.16] | *β* = -0.13<br>*t* = -1.65<br>*p* = 0.100<br>adj.*p* = 0.405<br>*CI* = [-0.29; 0.03] |
| | Log-win mtDNAcn | *β* = 2.95<br>*t* = 0.42<br>*p* = 0.678<br>adj.*p* = 0.916<br>*CI* = [-11.04; 16.94] | *β* = -0.06<br>*t* = -0.46<br>*p* = 0.645<br>adj.*p* = 0.914<br>*CI* = [-0.32; 0.20] | *β* = -0.10<br>*t* = -0.64<br>*p* = 0.525<br>adj.*p* = 0.857<br>*CI* = [-0.40; 0.21] |
| | MRI-BPAD | *β* = -0.41<br>*t* = -0.92<br>*p* = 0.357<br>adj.*p* =0.739<br>*CI* = [-1.30; 0.47] | *β* = -0.01<br>*t* = -1.12<br>*p* = 0.264<br>adj.*p* = 0.651<br>*CI* = [-0.02; 0.01] | *β* = 0.01<br>*t* = 1.44<br>*p* = 0.151<br>adj.*p* = 0.497<br>*CI* = [-0.01; 0.03] |
| | HorvathAA | *β* = -0.31<br>*t* = -0.82<br>*p* = 0.415<br>adj.*p* = 0.796<br>*CI* = [-1.07; 0.44] | *β* = -0.003<br>*t* = -0.46<br>*p* = 0.646<br>adj.*p* = 0.914<br>*CI* = [-0.02; 0.01] | *β* = 0.01<br>*t* = 0.58<br>*p* = 0.563<br>adj.*p* = 0.882<br>*CI* = [-0.01; 0.02] |
| | HannumAA | *β* = 0.19<br>*t* = 1.02<br>*p* = 0.307<br>adj.*p* = 0.679<br>*CI* = [-0.17; 0.55] | *β* = -0.001<br>*t* = -0.34<br>*p* = 0.738<br>adj.*p* = 0.933<br>*CI* = [-0.01; 0.01] | *β* = 0.00<br>*t* = 0.04<br>*p* = 0.969<br>adj.*p* = 0.987<br>*CI* = [-0.01; 0.01] |

| | | | |
|---|---|---|---|
| PhenoAA | $\beta = -0.06$ | $\beta = -0.003$ | $\beta = 0.002$ |
| | $t = -0.19$ | $t = -0.43$ | $t = 0.31$ |
| | $p = 0.847$ | $p = 0.668$ | $p = 0.757$ |
| | adj.$p = 0.953$ | adj.$p = 0.914$ | adj.$p = 0.933$ |
| | $CI = [-0.67; 0.55]$ | $CI = [-0.01; 0.01]$ | $CI = [-0.01; 0.02]$ |
| Horvath IEAA | $\beta = -0.43$ | $\beta = -0.01$ | $\beta = 0.01$ |
| | $t = -0.95$ | $t = -0.65$ | $t = 0.60$ |
| | $p = 0.343$ | $p = 0.518$ | $p = 0.549$ |
| | adj.$p = 0.730$ | adj.$p = 0.850$ | adj.$p = 0.871$ |
| | $CI = [-1.31; 0.46]$ | $CI = [-0.02; 0.01]$ | $CI = [-0.01; 0.03]$ |
| Hannum IEAA | $\beta = 0.60$ | $\beta = -0.004$ | $\beta = -0.01$ |
| | $t = 1.25$ | $t = -0.54$ | $t = -0.57$ |
| | $p = 0.211$ | $p = 0.589$ | $p = 0.571$ |
| | adj.$p = 0.574$ | adj.$p = 0.890$ | adj.$p = 0.883$ |
| | $CI = [-0.34; 1.55]$ | $CI = [-0.02; 0.01]$ | $CI = [-0.03; 0.01]$ |

**Abbreviations:** Absolute telomere length (aTL), age acceleration (AA), beta coefficient (*β*), Childhood Trauma Questionnaire-Short Form (CTQ-SF), extrinsic epigenetic age acceleration (EEAA), FDR adjusted *p*-value (adj.*p*), Human Immunodeficiency Virus (HIV), intrinsic epigenetic age acceleration (IEAA), log-winsorised (Log-win), magnetic resonance imaging-based brain-predicted age difference (MRI-BPAD), mitochondrial DNA copy number (mtDNAcn), *p*-value (*p*), t-statistic (*t*), 95% confidence interval (CI).

**Significance:** $^{*}\ p < 0.05$, $^{**}\ p < 0.01$, $^{***}\ p < 0.001$
